# Benchmarking Deep Learning-based Image Retrieval of Oral Tumor Histology

**DOI:** 10.1101/2024.05.30.24308181

**Authors:** Ranny Rahaningrum Herdiantoputri, Daisuke Komura, Mieko Ochi, Yuki Fukawa, Kou Kayamori, Maiko Tsuchiya, Yoshinao Kikuchi, Tetsuo Ushiku, Tohru Ikeda, Shumpei Ishikawa

## Abstract

Oral tumors necessitate a dependable computer-assisted pathological diagnosis system considering their rarity and diversity. A content-based image retrieval (CBIR) system using deep neural networks has been successfully devised for digital pathology. No CBIR system for oral pathology has been investigated because of the lack of an extensive image database and feature extractors tailored to oral pathology. This study uses a large CBIR database constructed from 30 categories of oral tumors to compare deep learning methods as feature extractors. The highest average area under the receiver operating curve (AUC) was achieved by models trained on database images using self-supervised learning (SSL) methods (0.900 with SimCLR; 0.897 with TiCo). The generalizability of the models was validated using query images from the same cases taken with smartphones. When smartphone images were tested as queries, both models yielded the highest mean AUC (0.871 with SimCLR and 0.857 with TiCo). We ensured the retrieved image result would be easily observed by evaluating the top-10 mean accuracy and checking for an exact diagnostic category and its differential diagnostic categories. Therefore, training deep learning models with SSL methods using image data specific to the target site is beneficial for CBIR tasks in oral tumor histology to obtain histologically meaningful results and high performance. This result provides insight into the effective development of a CBIR system to help improve the accuracy and speed of histopathology diagnosis and advance oral tumor research in the future.

## Introduction

Oral tumors are generally composed of diverse and rare tumor types, except for major categories like squamous cell carcinoma. Distinguishing oral tumor types is difficult except for well-experienced oral pathologists. The rarity of oral tumors and the diverse tissue types in the oral region make obtaining reference images for diagnosis and research a challenge, potentially leading to delayed diagnosis and a significant burden on pathologists [1]. Consequently, a diagnostic system is needed to improve the speed and accuracy of histopathological diagnosis of these tumors [2]. Artificial intelligence (AI) is a promising solution for efficient histopathological diagnosis of oral tumors.

AI development for oral tumor diagnosis is limited and focused only on a few tumor types. Classification methods have been developed to predict the diagnosis, such as ameloblastoma or odontogenic keratocysts, to which a histopathological image may belong [3,4]. These approaches are helpful in common cases. However, a computer-aided diagnostic system that covers a broader spectrum of tumor types would be more practical and would help narrow the differential diagnoses. Therefore, content-based image retrieval (CBIR) is suitable. CBIR regards histopathological images as query images to find similar images from a database based on their similar morphology [2,5]. This system is useful as a diagnostic aid for finding case references, especially where diagnostic expertise is challenging to find, such as in low-to middle-income countries [1]. The involvement of human intervention is crucial in diagnosis. Conventionally, pathologists diagnose directly after H&E-stained slide analysis or optionally use different methods as diagnostic aids: referring atlases, consulting subspecialist experts, or conducting ancillary tests. An automatic image search can complement these options to expedite image reference search (Figure 1). With scarce pathological expertise, a tool that could provide urgently needed information for rapid diagnosis before conducting tests to raise a definitive one would be significant [6]. CBIR provides interpretability because it presents multiple candidate images, which is beneficial when distinguishing between categories based on histopathological images alone is challenging, such as when information on dental infections or radiographic findings is needed. With CBIR, the retrieved results are to be evaluated by pathologists, reducing the risk of misdiagnosis owing to inaccurate results, especially for categories with very similar histology.

**Figure 1:**
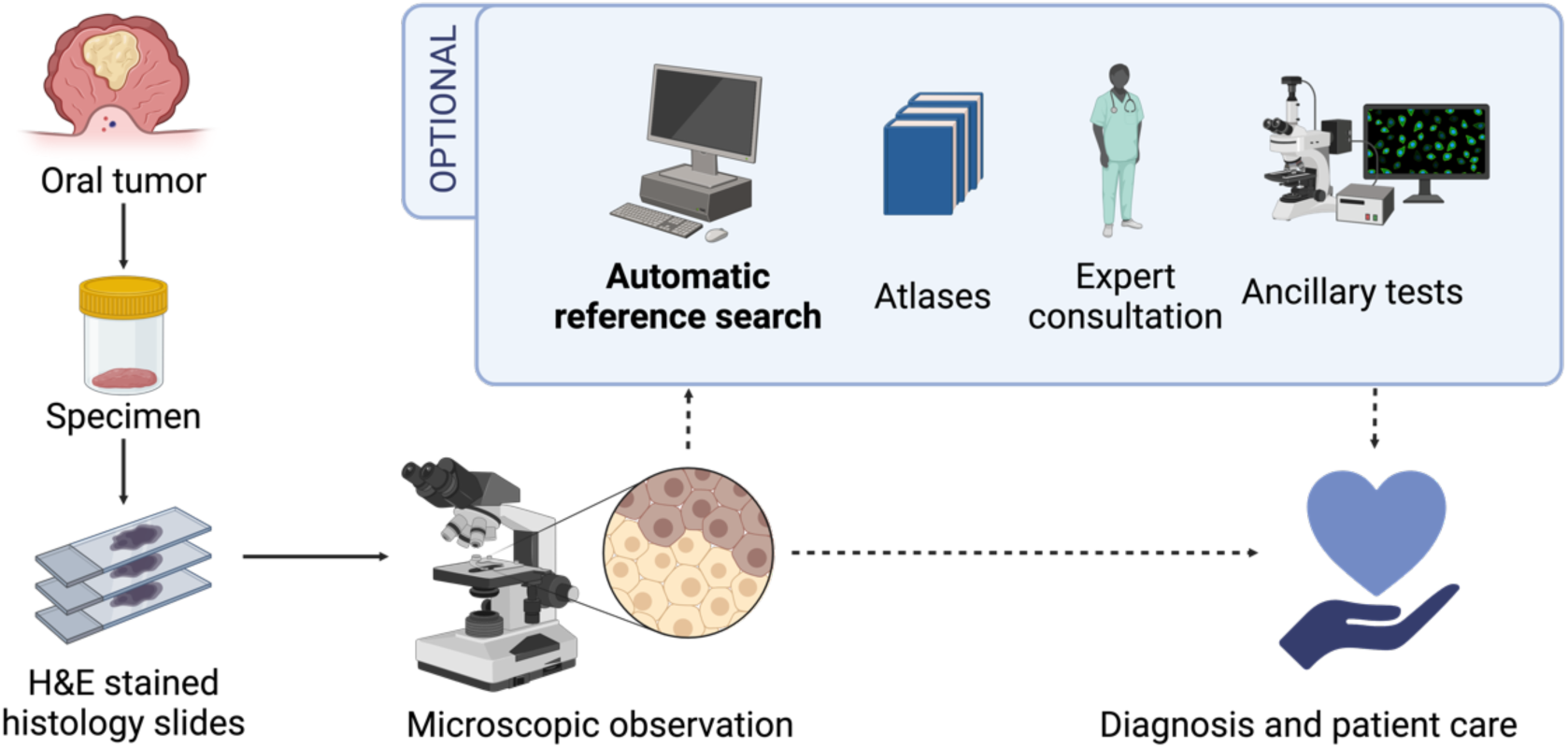
CBIR’s place in the pathological diagnosis workflow. CBIR optionally providing an interpretable automatic reference search that is fast and easily followed up with more thorough study with atlases, discussions, or ancillary tests. CBIR could help point out similar features from the previously diagnosed image in the database that may lead to testable differential diagnoses more swiftly than directly consulting atlases or senior experts, which may cause the patient delayed treatment. (Image created with BioRender)

The CBIR system consists of two aspects: image feature extraction and nearest-neighbor search. Feature extraction is crucial because it must adequately capture complex histological features such as staining patterns, tissue structures, and cellular morphology to create histologically relevant image representation [2,7,8]. The extracted features must be robust to irrelevant color variations, such as different hematoxylin and eosin (HE) stain brands, glass slide color degradation, and image-capturing devices ranging from whole-slide image (WSI) scanners to smartphone cameras [5,8,9]. At the early stage of CBIR development, traditional image features such as shape, color, texture, or a combination were used. Recent developments showed that deep learning models outperformed traditional features [6,7]. Several deep learning methods, such as supervised learning where models are pre-trained on general images or fine-tuned on histopathological images, have been used to train feature extractors [9–13], and self-supervised learning (SSL), which allows learning from unlabeled images [14–16]. However, no studies have reviewed which method is most suitable for CBIR in oral tumors.

This study aimed to investigate the performance of different deep learning models for oral tumor CBIRs by developing a large dataset of whole-slide images from 541 cases with 51 tumor types and evaluating the retrieval accuracy by comparing different representational learning techniques.

## Materials & Methods

### Dataset

We collected diagnostic slides of the oral tumor categories described in Chapters 7 and 8 of the WHO Classification of Head and Neck Tumors, 4^th^ Edition [17]. Patients were diagnosed in 2001–2022 and underwent surgery at Tokyo Medical and Dental University (TMDU) Hospital. Patients or their surrogates had the option to withdraw from this study through public notices according to the approved protocols. This study was approved by the Institutional Review Board (IRB) of TMDU (No. D2019-087). Some slides that were lost, broken, or required diagnostic confirmation with immunohistochemistry (IHC) staining were remade from the paraffin-embedded tissue blocks. Additional IHC staining was done for the secretory carcinoma and the atypical acinic cell carcinoma cases older than 2017. Categories with fewer than five cases were excluded. All slides that fulfill the inclusion criteria were scanned using a NanoZoomer S210 slide scanner (C13239-01; Hamamatsu Photonics, Japan) at 40× magnification. The tumor areas were annotated by a pathology resident and verified by board-certified pathologists. We included the tumor areas that are typical to the tumor category while excluding the normal tissue and the severe artifacts such as torn or folded tissue. Image patches were then randomly extracted from the annotated tumor areas with three different sizes: 905 µm, 453 µm, and 226 µm. Twenty image patches were extracted with each magnification. The use of three different sizes was to accommodate different magnification levels and preserve the histologic information at the tissue and cellular level as much as possible. The dataset comprises 49,243 image patches from 51 categories, covering approximately 50% of the oral tumor categories (Table 1).

**Table 1.**
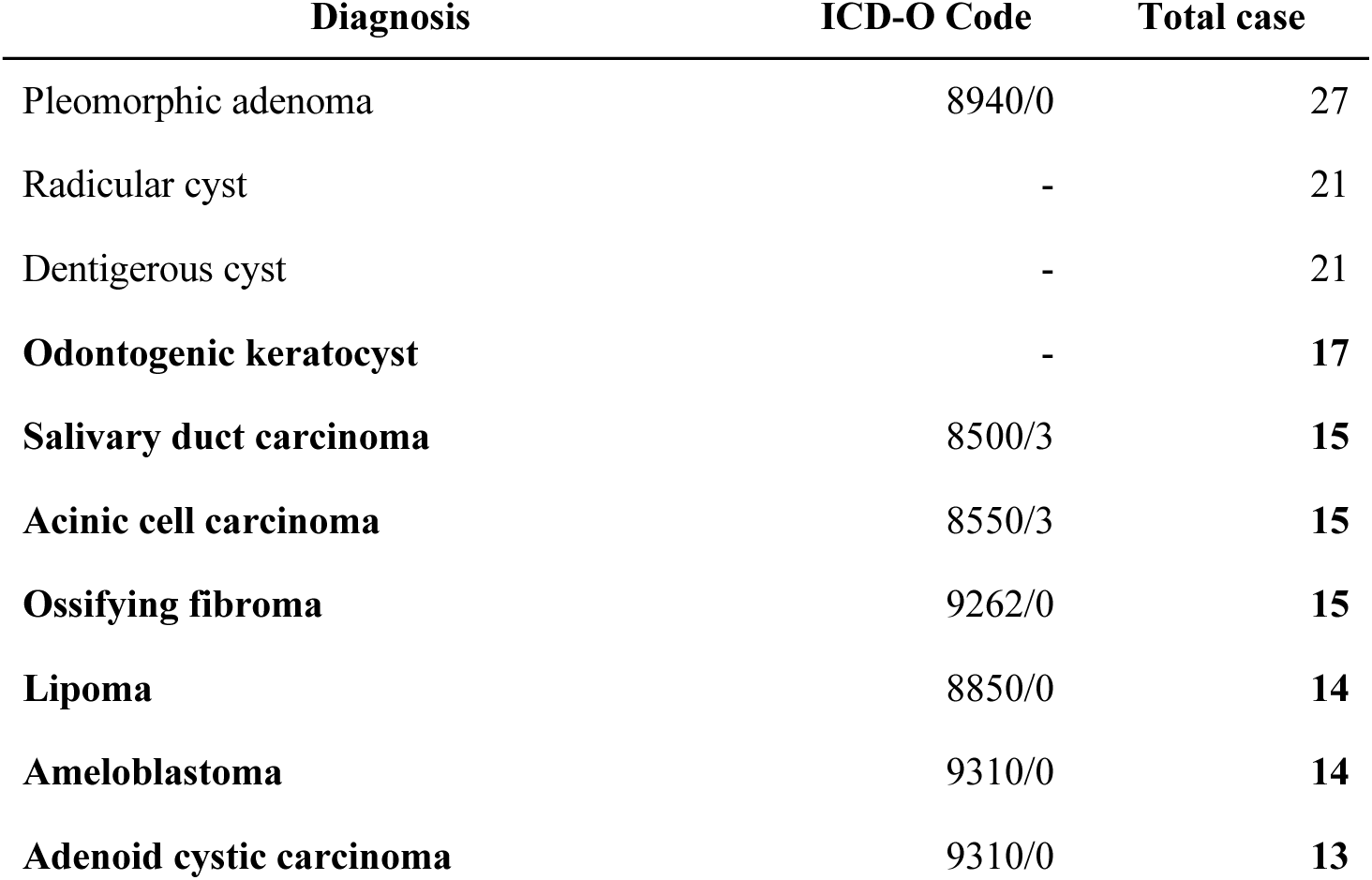
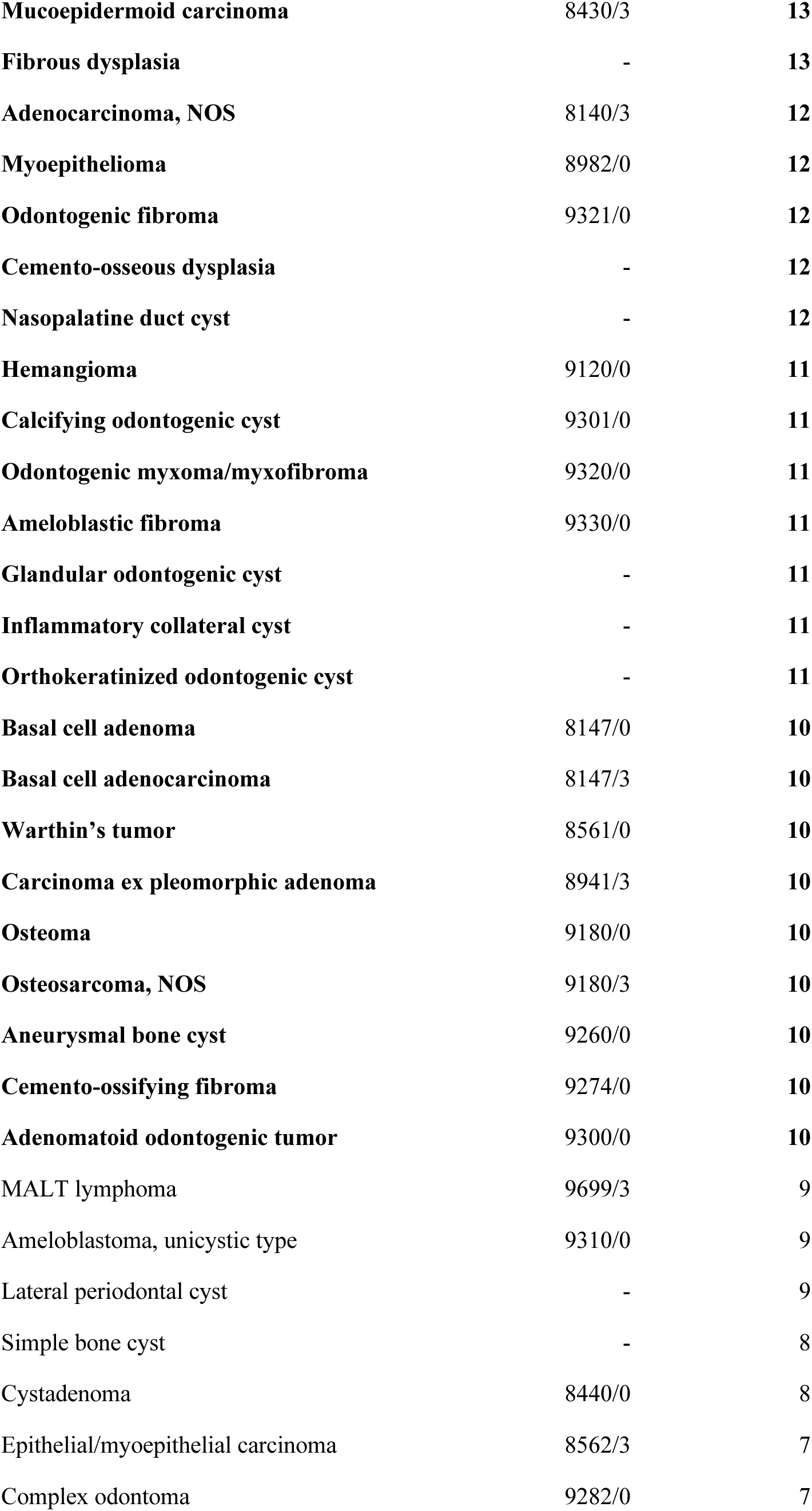
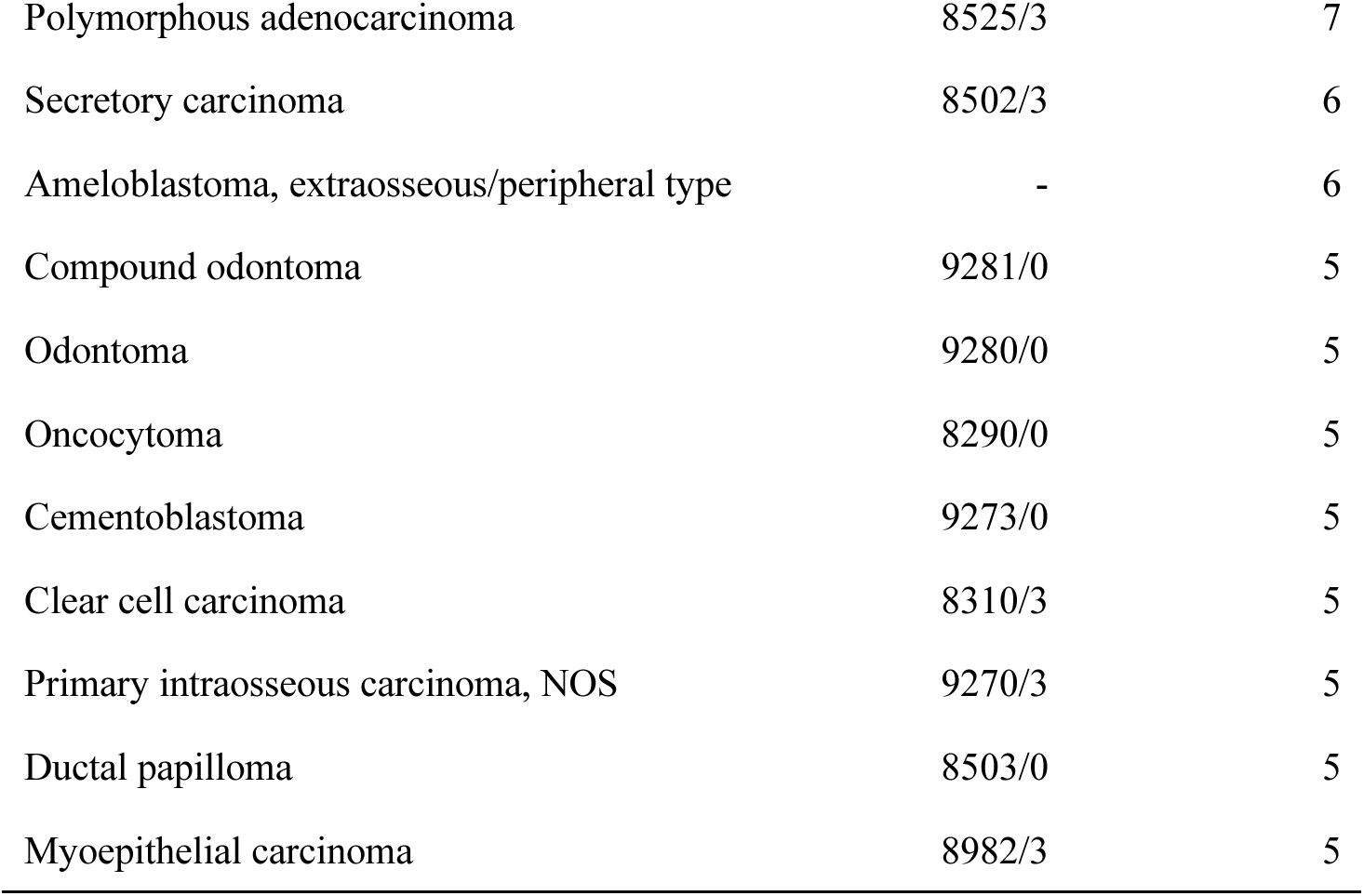
The tumor categories included in the dataset, their corresponding ICD-O codes, and the total number of cases. Some categories do not correspond to the ICD-O but are described in the WHO Classification of Head and Neck Tumors. 4th Ed. Categories consisting of ten to twenty cases, marked in **bold**, were included in the CBIR database.

### Database construction

A database from a subset of the dataset containing at least ten cases was compiled. Image representations from each model’s encoder were stored in the database (Figure 2A). It contains 33,356 image patches from 30 oral tumor categories (Table 1).

**Figure 2:**
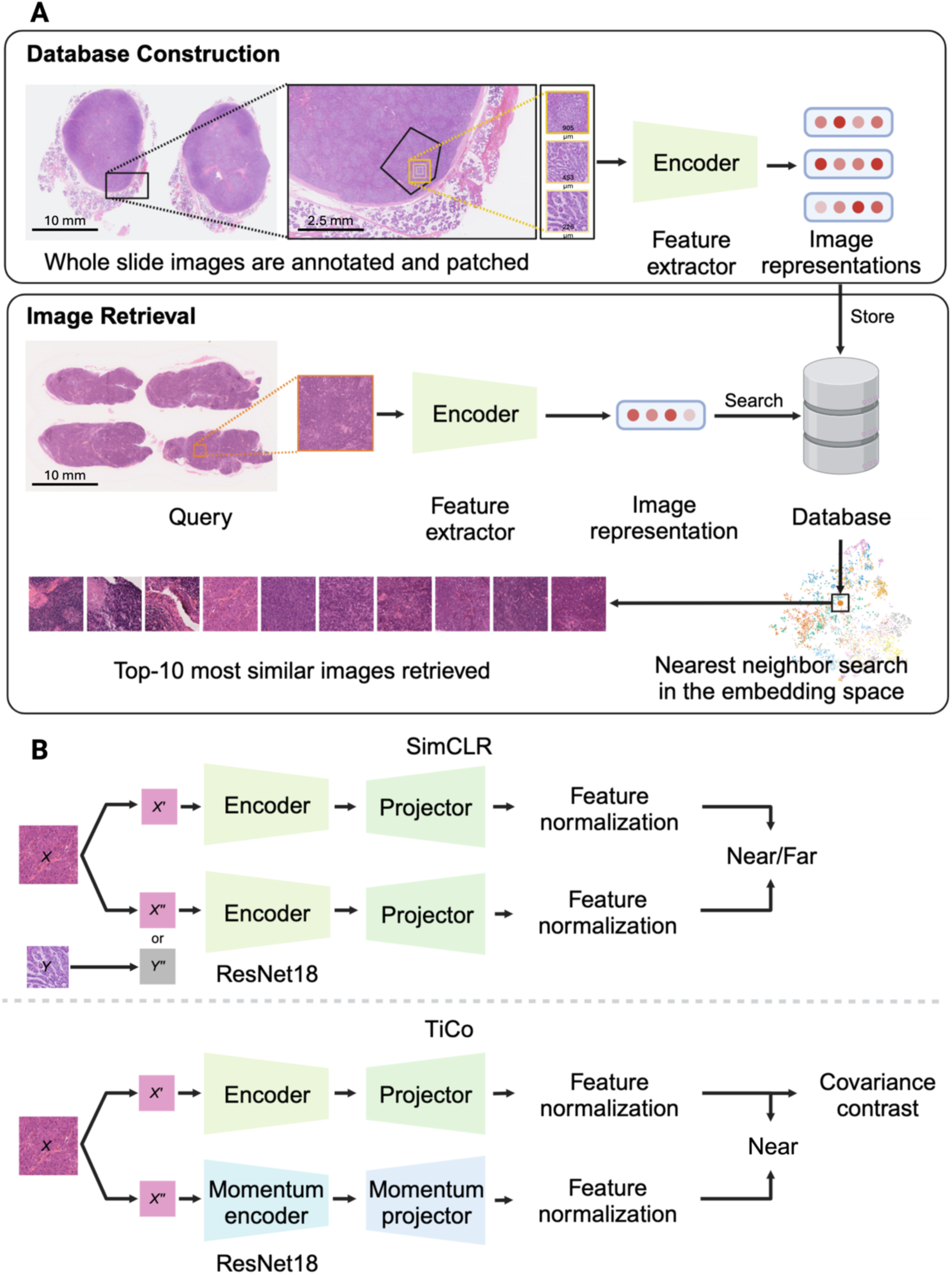
CBIR workflow and SSL models training process. (A) Schematic representation of CBIR using deep neural networks to retrieve similar oral tumor histopathological images. The similarity is determined by a nearest-neighbor search, which calculates the cosine similarity of the query image’s image representation to all database image representations in the embedding space. (B) The training process of the SSL models used ResNet18 as an encoder. The image representations were passed to a projector and subjected to feature normalization. In the SimCLR method, the training loss function yields a low value when the representation of the original image (X′) and its augmentation (X′′) are close together, while it yields a larger value when X′ and a different image augmentation (Y′′) are far apart. In the TiCo method, the process still pulls X′ and X′′ close, and then the redundancy in the representation is removed using covariance contrast without using a different image (Y′′). (Image created with BioRender)

### Test queries

The test queries were the cases available in the hospital repository after the collection of the database case was finished and were representatives of the tumor major categories in the database: odontogenic cysts, odontogenic tumors, benign and malignant tumors of the salivary gland, maxillofacial bone tumor, and soft tissue tumor. Slides that include severe artifacts that are impossible to avoid when extracting image patches were excluded. We prepared three query sets from different hospitals to test the performance: Query case set-A was collected between 2022 and 2023 from the repository of the same institution (TMDU) as the database. Only the tumor categories that have two representative cases were included as the test queries. Finally, eleven tumor categories with two cases in each were used as the test queries. Histopathologic slides were scanned to create WSIs for in-domain queries with the same device as the database image. Three selected tumor areas that are typical of the tumor type from the same slides were photographed with smartphone cameras (Samsung S21FE, iPhone 6, and Motorola g8) using an Olympus BX53 microscope with 10×, 20×, and 40× objective lens magnification to create out-of-domain-phonecam queries. The location (indoor laboratory environment) and the amount of light from the microscope were unchanged when taking the smartphone images. The query case set-B was compiled from the University of Tokyo case (approved by the IRB of The University of Tokyo No. 2019158NI). Eleven cases from eight salivary gland tumor categories were included in this study. Only benign and malignant tumors of the salivary gland could be collected for out-of-domain-B since no other major categories exist in the repository. Histopathologic slides were scanned using a NanoZoomer 2.0HT slide scanner (C9600-12, Hamamatsu Photonics, Japan) to create WSIs for out-of-domain-B queries. The query case set-C was collected from Teikyo University Hospital from 2018-2023 (approved by the IRB of Teikyo University No. 23-054). Only the tumor categories that have two representative cases were included as the test queries. Twenty cases from eight oral tumor categories were included in the study. The histopathologic slides were scanned using a NanoZoomer XR slide scanner (C12000-02; Hamamatsu Photonics, Japan) to create WSIs for out-of-domain-C queries. Patients for query cases set-B and set-C or their surrogates had the option to withdraw from this study through public disclosure according to the approved protocols.

For the WSI queries, the same method was used to create image patches from the WSI as from the database image. All scanned WSIs were annotated to 3–4 representative tumor areas per WSI. From these areas, 20 image patches per magnification level (905 µm, 453 µm, and 226 µm) per slide were extracted to create image patches. The total number of image patches used for the evaluation was 2,520 images from the WSI and 594 smartphone images.

The representation of each query image was calculated with each tested model. The nearest-neighbor search was performed based on cosine similarity with the database images. Examples of query images for each category in each set can be found in Figures 3–5. The detailed methods for database construction, including the tumor areas selection, patch extraction, feature extraction code, and image retrieval were adapted from our previous study [18].

**Figure 3:**
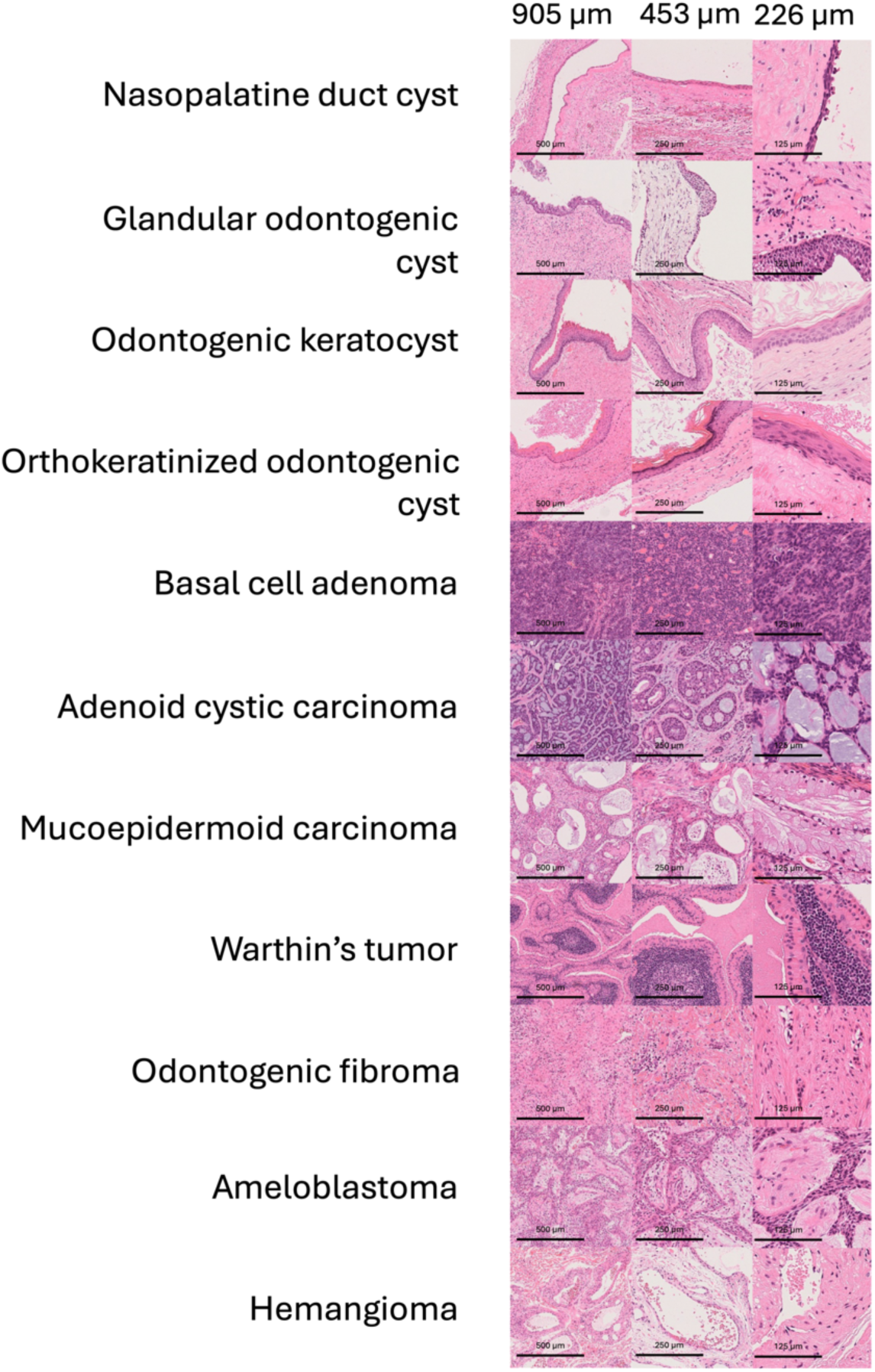
Examples of set-A query images from each category for each magnification level. The total number of image patches extracted from WSIs is 1,320 images.

**Figure 4:**
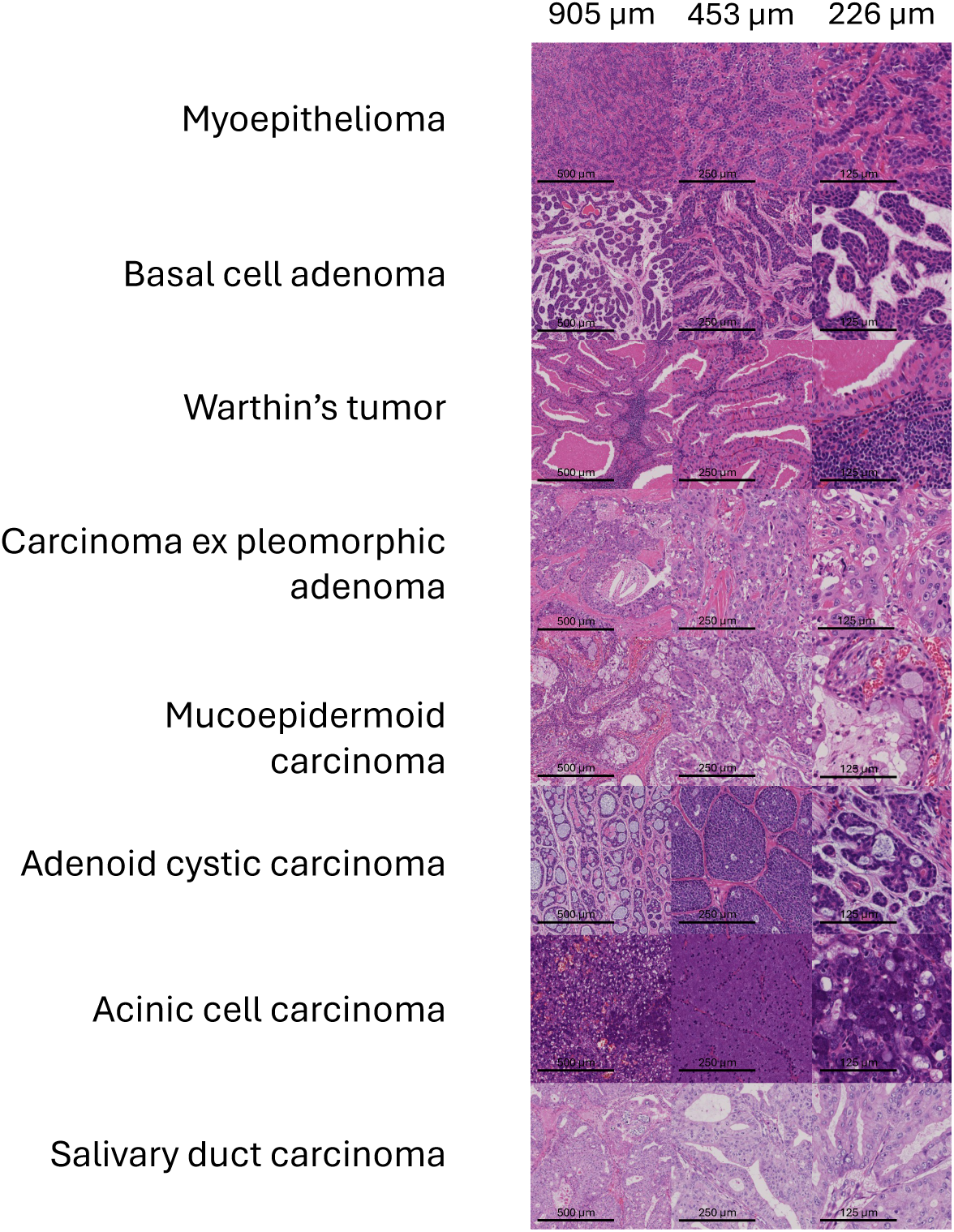
Examples of set-B query images from each category for each magnification level. The total number of image patches extracted from the WSIs is 660 images.

**Figure 5:**
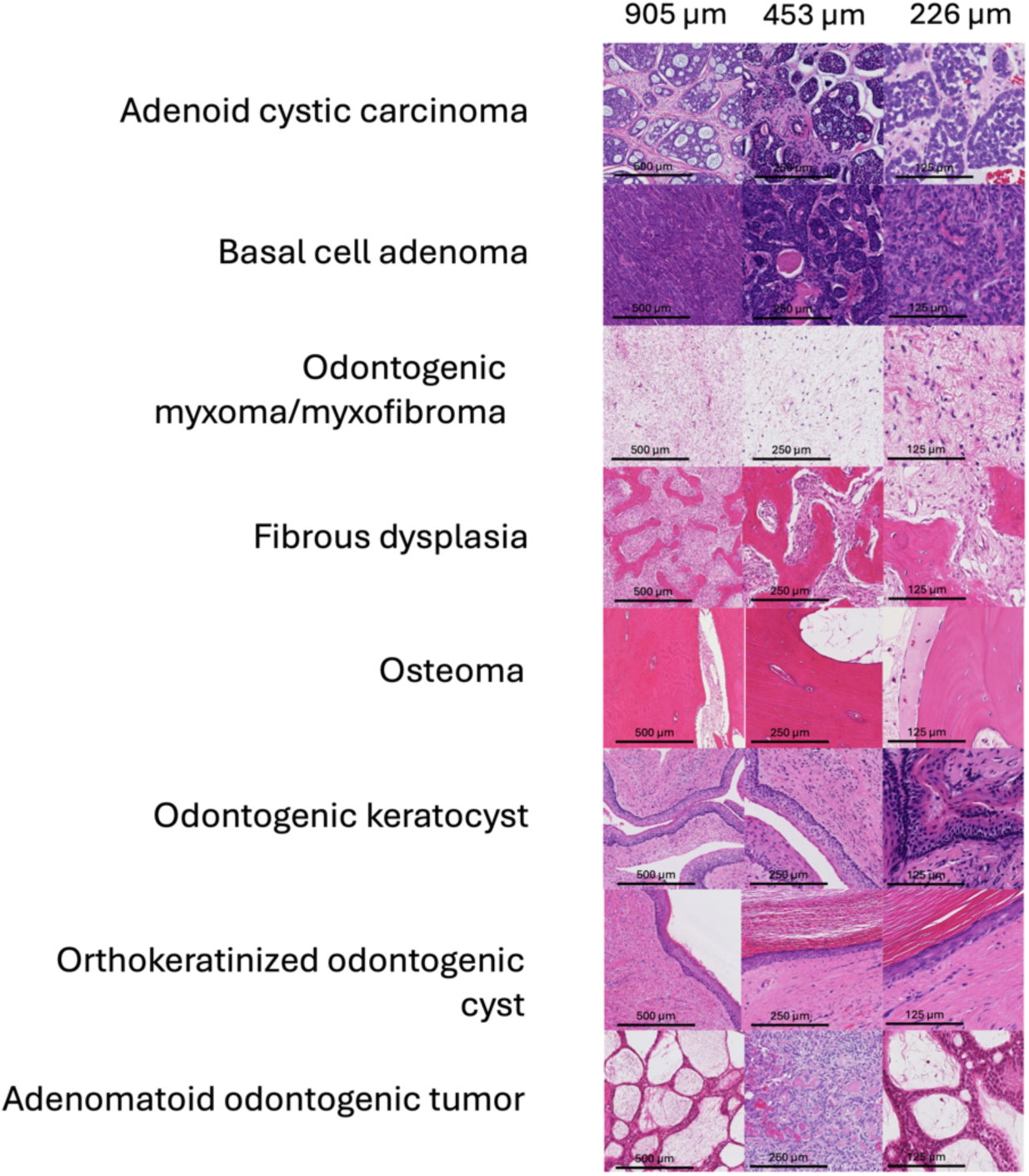
Examples of set-C query images from each category for each magnification level. The total number of image patches extracted from the WSIs is 1,200 images.

### Evaluation metrics and statistical analysis

The area under the receiver operating characteristic curve (AUC) for all query images with top-*k* retrieved images (*k* ranges from 1 to the total number of cases in the database) being the cut-points were averaged into MeanAUC. Based on the top-10 images most similar to the query, three additional metrics were evaluated. MeanAcc denotes the mean of the top-10 diagnostic accuracies (Acc) for each query. %query denotes the percentage of results that contained at least one accurate diagnosis category. The histological similarity in the retrieved results beyond diagnostic accuracy was evaluated by noting the retrieved images that did not belong to the accurate diagnosis category or any of its differential diagnoses [17,19,20] (Table 2). The values are expressed as histologic inaccuracy (HI) and were averaged to determine the MeanHI. Image retrieval and all statistical analyses were conducted using Python 3.7.12 and R 4.2.2.

**Table 2.** List of test query categories and all their respective differential diagnoses that are represented in the database for MeanHI evaluation.

| Query Category |  | Differential Diagnoses |  |
| --- | --- | --- | --- |
| Acinic cell carcinoma | Salivary duct carcinoma | Secretory carcinoma | Mucoepidermoid carcinoma |
| Adenoid cystic carcinoma | Basal cell adenocarcinoma | Basal cell adenoma |  |
| Adenomatoid odontogenic tumor | Ameloblastoma | Mucoepidermoid carcinoma |  |
| Ameloblastoma | Adenomatoid odontogenic tumor | Ameloblastic fibroma |  |
| Basal cell adenoma | Adenoid cystic carcinoma | Basal cell adenocarcinoma | Myoepithelioma |
| Carcinoma ex pleomorphic adenoma | Salivary duct carcinoma | Adenocarcinoma, NOS | Mucoepidermoid carcinoma |
| Fibrous dysplasia | Ossifying fibroma | Osteosarcoma, NOS |  |
| Glandular odontogenic cyst | Nasopalatine duct cyst | Inflammatory collateral cyst | Mucoepidermoid carcinoma |
| Hemangioma | Ossifying fibroma | Aneurysmal bone cyst |  |
| Mucoepidermoid carcinoma | Adenocarcinoma, NOS | Warthin's tumor | Carcinoma ex pleomorphic adenoma |
| Myoepithelioma | Mucoepidermoid carcinoma | Basal cell adenoma |  |
| Nasopalatine duct cyst | Glandular odontogenic cyst | Inflammatory collateral cyst |  |
| Odontogenic fibroma | Odontogenic myxoma/myxofibroma | Ossifying fibroma |  |
| Odontogenic keratocyst | Orthokeratinized odontogenic cyst | Ameloblastoma |  |
| Odontogenic myxoma/myxofibroma | Odontogenic fibroma |  |  |
| Orthokeratinized odontogenic cyst | Odontogenic keratocyst | Ameloblastoma |  |
| Osteoma | Ossifying fibroma | Fibrous dysplasia |  |
| Salivary duct carcinoma | Acinic cell carcinoma | Secretory carcinoma | Adenocarcinoma, NOS |
| Warthin's tumor | Mucoepidermoid carcinoma |  |  |

### Model preparation and training

#### ImageNet-1k Pre-trained CNN

We tested VGG16 pre-trained on 1.2 million images from ImageNet after it was shown to perform well as an image classifier in several studies [22,23]. In this study, the block4_conv3 layer was used as the feature extractor because the middle layer of a convolutional neural network (CNN) architecture has been shown to capture features that are more suitable for histopathology images [12].

#### ImageNet-22k Pre-trained Vision Transformer

We used the DINOv2 ViT-L/14 model pre-trained with the SSL method on general images from ImageNet-22k [26]. The last layer was used for feature extraction. The images were cropped to 252 pixels owing to the input size restrictions.

#### Fine-tuned CNN

We fine-tuned all the ImageNet-1k pre-trained ResNet18 models on our dataset using the supervised learning method to classify 51 categories. All the layers were trained with a learning rate of 0.001, a batch size of 32, and 100 epochs in PyTorch 1.11.0. During training, a random 90-degree rotation, random horizontal and vertical flips, color jitter, Gaussian blur, and color normalization transformation were performed. The training-to-test ratio was 8:2.

#### CNN Trained with SSL Methods

Contrastive (SimCLR) [21] and noncontrastive methods (TiCo) [25] were investigated. ResNet18 is used as the backbone. During training, random color jittering, grayscale, image scaling, horizontal and vertical flipping, 90-degree rotation, Gaussian blurring, and color augmentation were implemented (Figure 6). Both models were trained with Lightly version 1.3.3, with a learning rate of 1.2, a batch size of 32 × 32 (with accumulated gradients), and 1,000 epochs. Examples of original and augmented images are shown in Figure 2B. The ResNet18 backbone model trained on 57 histopathology image datasets (38,594 image patches and 24,923 WSIs) developed by Ciga et al. 2022 was also used for comparison [14].

**Figure 6:**
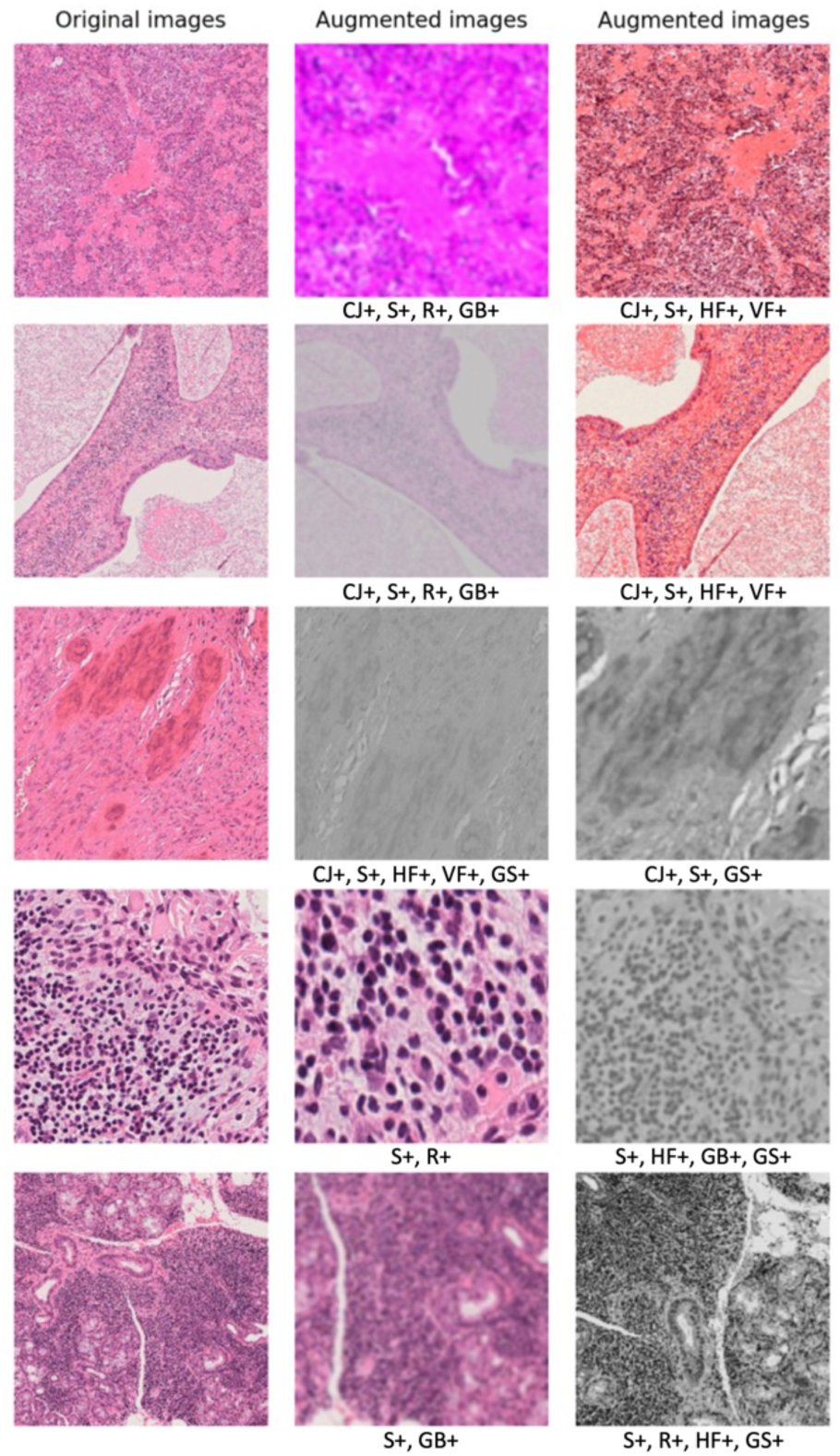
Data augmentation examples. Random color jittering (CJ+), grayscale (GS+), image scaling (S+), horizontal and vertical flips (HF+ and VF+), 90-degree rotation (R+), and Gaussian blur (GB+), used during SSL training. Color normalization was used during model training but is not included in this visualization.

The code for SSL model training is available at https://github.com/rannyrh/oralpath_CBIR.

#### Histopathology Image-trained Vision Transformer

A vision transformer-based model, Phikon, was trained with 40 million pan-cancer tiles extracted from The Cancer Genomic Atlas (TCGA) using the masked image modeling (MIM) method as an SSL framework. MIM learns meaningful representation by randomly masks portions of an image and trying to reconstruct those masked portions. This model was developed by Owkin. Inc [24]

## Results

### Model performance evaluation

The highest MeanAUC for in-domain queries was achieved by ResNet18+SimCLR (0.900), followed by ResNet18+TiCo (0.897). They achieved this at the query category level for 8 out of the 11 categories. The generalizability of these models was validated using out-of-domain-phonecam queries. The highest performance for out-of-domain-phonecam queries was also achieved by ResNet18+SimCLR (0.871), followed by ResNet18+TiCo (0.857). The highest performance on the query case category levels was achieved by both SSL models for 7 out of the 11 categories. We tested the performance on queries from other institutions to further demonstrate the generalizability. A similar result was yielded by both SSL models, where ResNet18+SimCLR leads with the highest MeanAUC (0.886 for out-of-domain-B; and 0.913 for out-of-domain-D queries), followed by ResNet18+TiCo (0.881 for out-of-domain-B; and 0.905 for out-of-domain-D queries). Phikon, pre-trained on histopathological images, and DINOv2, pre-trained on large-scale general images, performed comparably well with DINOv2 leading the overall MeanAUC in out-of-domain-phonecam query, with Phikon leading in the other three query sets (Table 3).

**Table 3:**
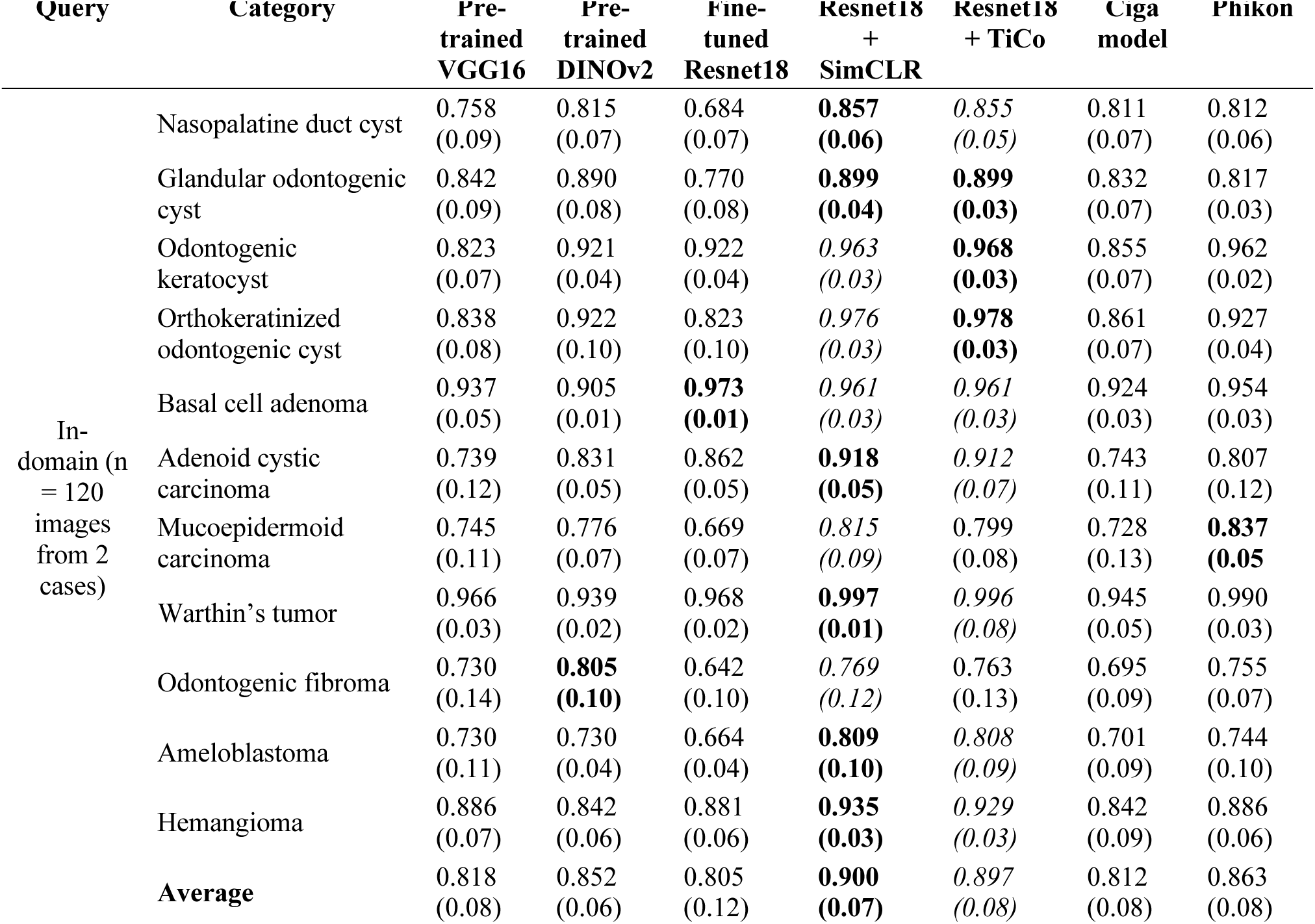
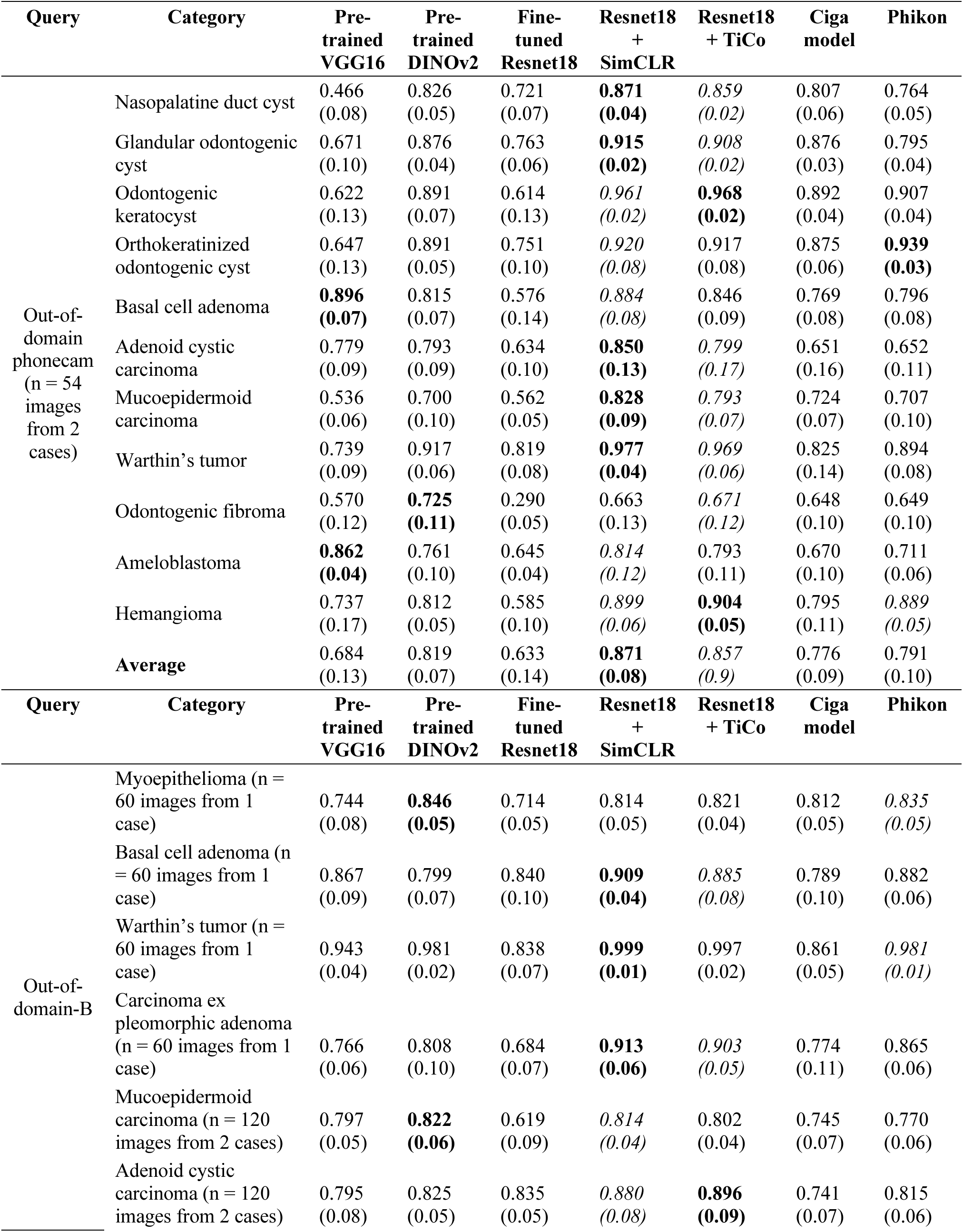
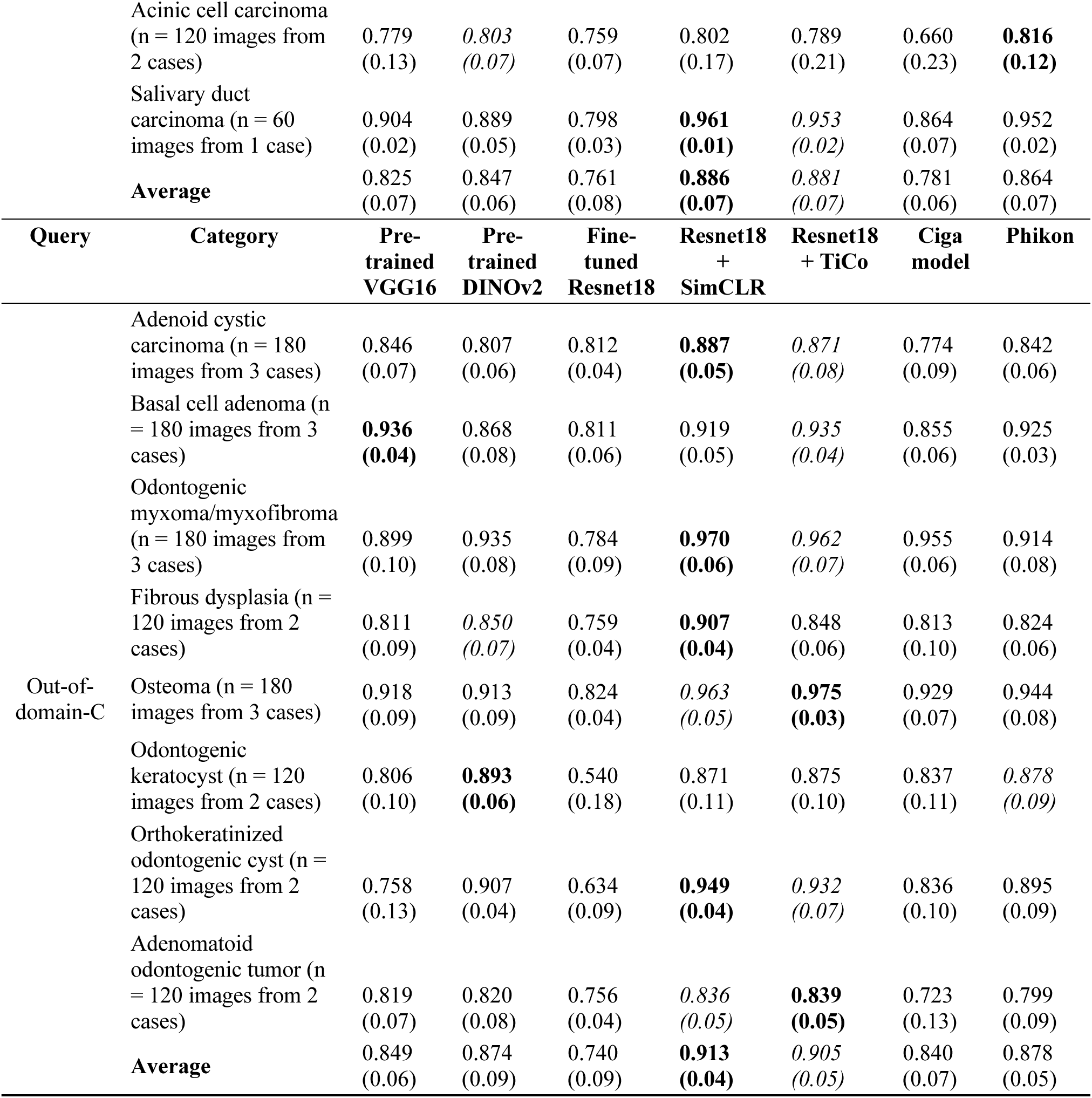
Mean-AUC (SD) of each test query category in in- and out-of-domain image queries. The performances of the SSL models are superior for most test query categories and the overall averages. The highest AUC for each category is marked in **bold**, and the second highest is marked in *italics*.

Overall, the MeanAcc of test query set-A was highest with the SSL models: ResNet18+TiCo outperformed other models for in-domain queries (4.64), followed by ResNet18+SimCLR (4.53) with no significant difference (Wilcoxon signed-rank test with Bonferroni adjustment). The reverse was observed for the out-domain-phonecam queries (ResNet18+SimCLR (3.33) and ResNet18+TiCo (3.31)) with no significant difference (Wilcoxon signed-rank test with Bonferroni adjustment). Phikon yielded the highest MeanAcc for out-of-domain-B queries (3.79), followed by ResNet18+SimCLR (3.68). Pre-trained DINOv2 outperformed other models for out-of-domain-C queries (3.64), followed closely by ResNet18+TiCo (3.63) (Table 4). The highest overall MeanAcc was consistently achieved by the SSL models at different magnification levels, except for the high-magnification in-domain queries (Figures 7A, 7B). The highest %query was obtained with SSL models for most query categories (Table 5).

**Figure 7:**
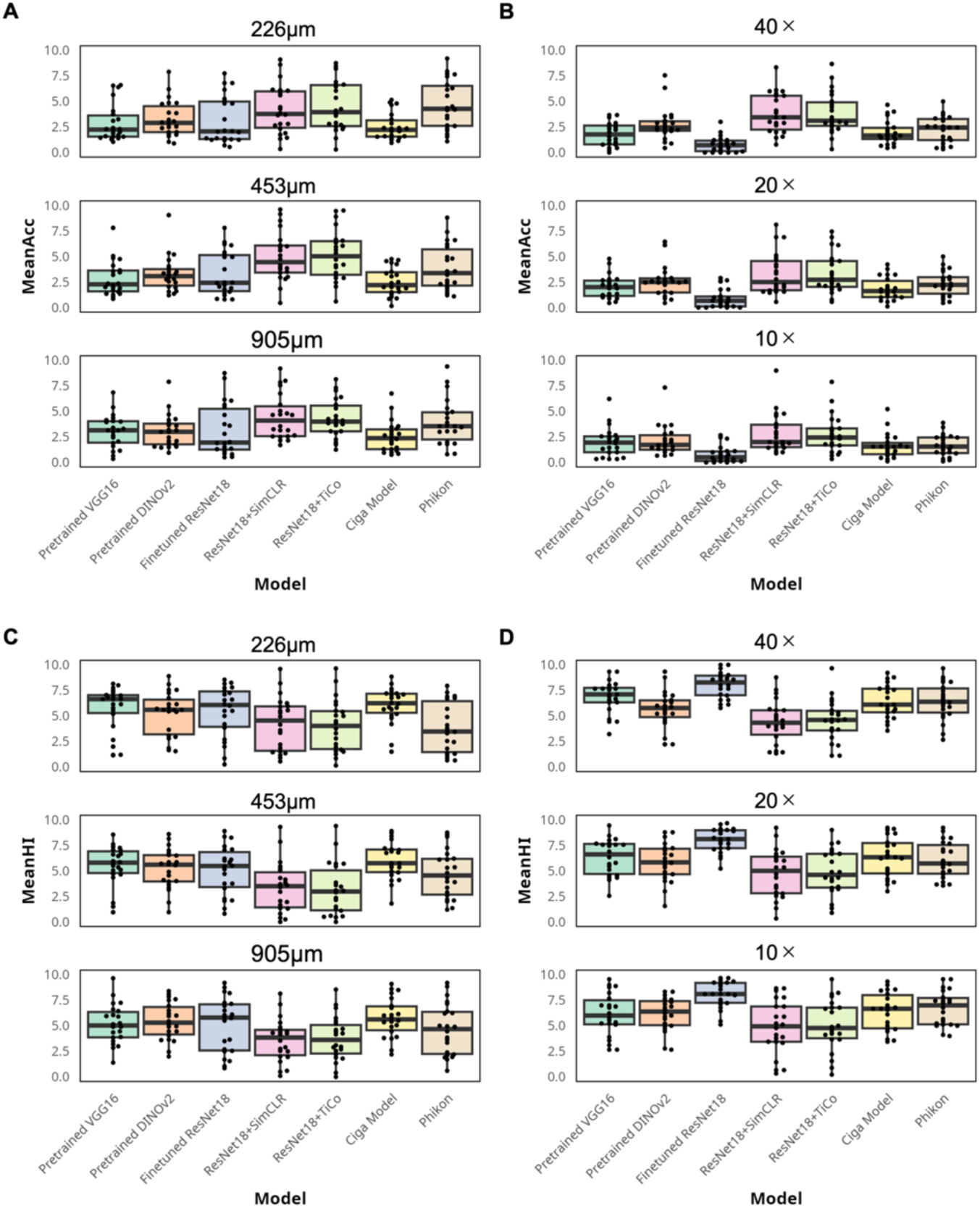
MeanAcc and MeanHI comparisons. (A) MeanAcc comparison for the in-domain query of all models by magnification showing the highest performance of Phikon at the highest magnification and that of ResNet18+SimCLR and ResNet18+TiCo at the moderate and lowest magnification. (B) MeanAcc comparison for out-of-domain-phonecam queries of all models by magnification shows the highest performance of ResNet18+SimCLR and ResNet18+TiCo at the highest and lowest magnification. Both model performances were comparable to that of pre-trained DINOv2 at moderate magnification. (C) MeanHI comparison for in-domain queries by magnification shows that ResNet18+SimCLR and ResNet18+TiCo outperformed other models except for the highest magnification where Phikon leads with a wider interquartile range. (D) MeanHI comparison for out-of-domain-phonecam query by magnification showing ResNet18+SimCLR and ResNet18+TiCo outperformed other models. (C-D) Please note that a lower MeanHI value denotes a higher model performance.

**Table 4.**
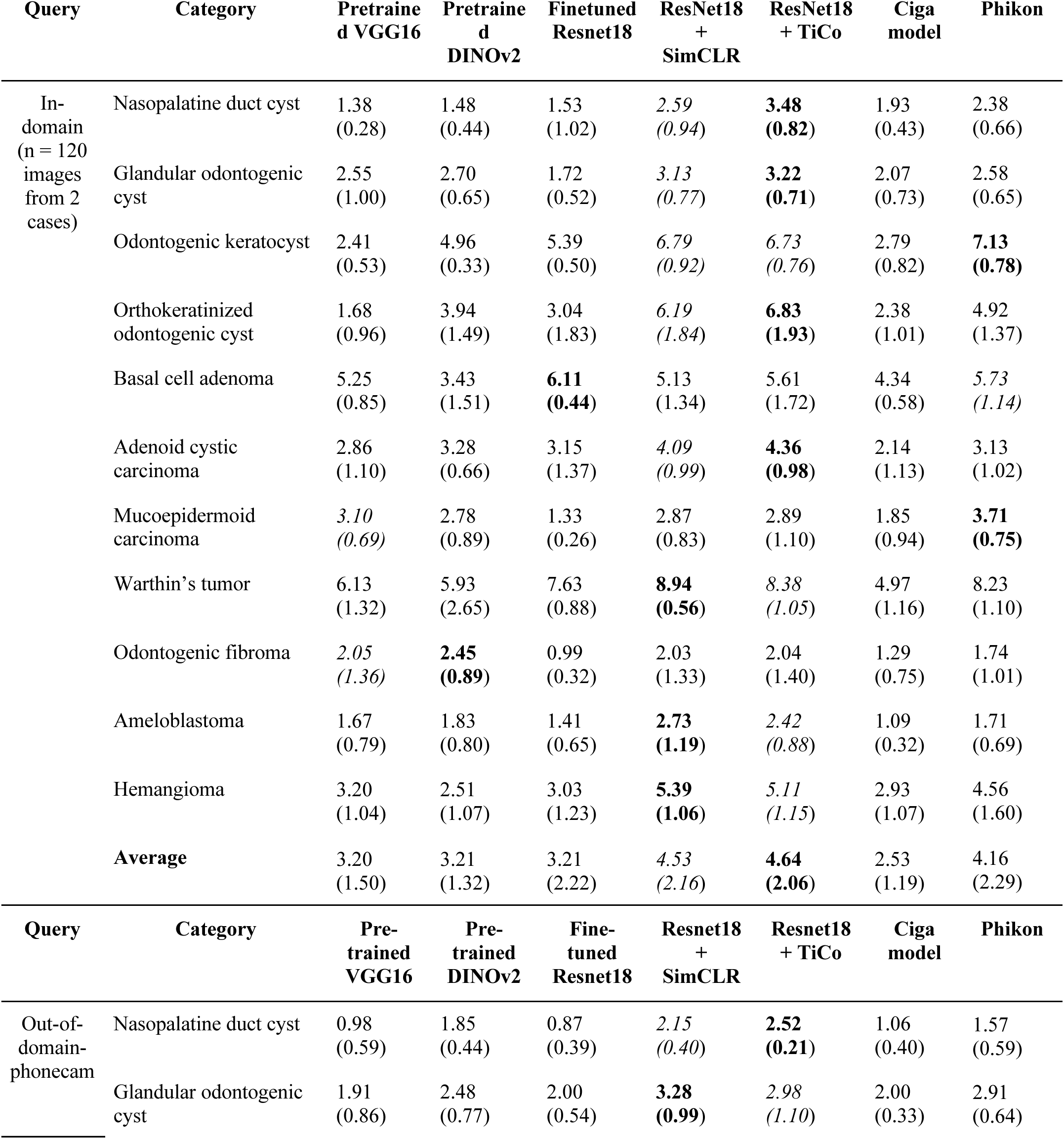
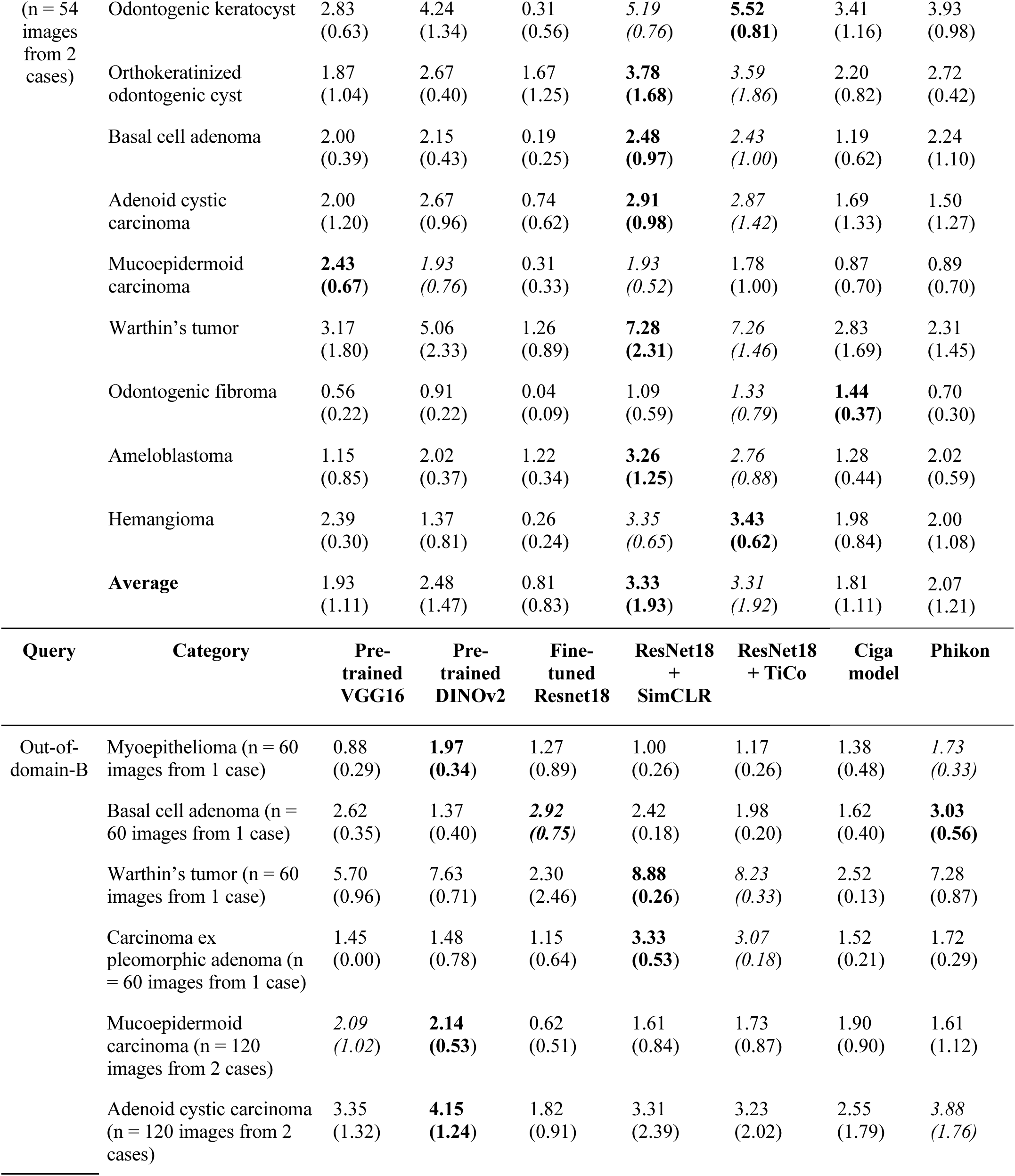
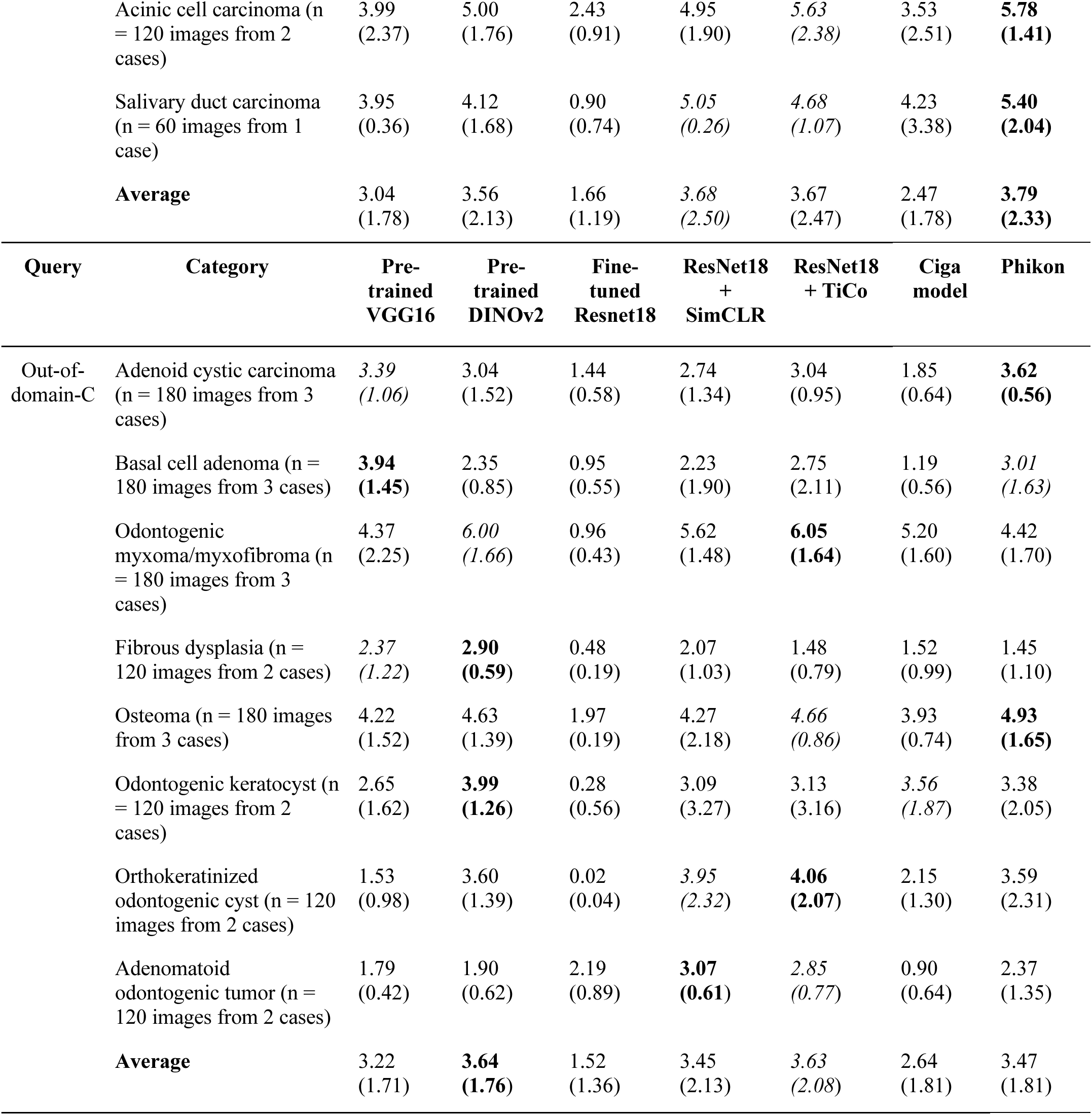
MeanAcc (SD) of each test query category in in- and out-of-domain image queries. The performances of the SSL models are superior for most categories, further validating the robustness of the models under a wide range of histopathological image conditions. The highest MeanAcc for each category is marked in **bold** and the second highest is in *italics*.

**Table 5.**
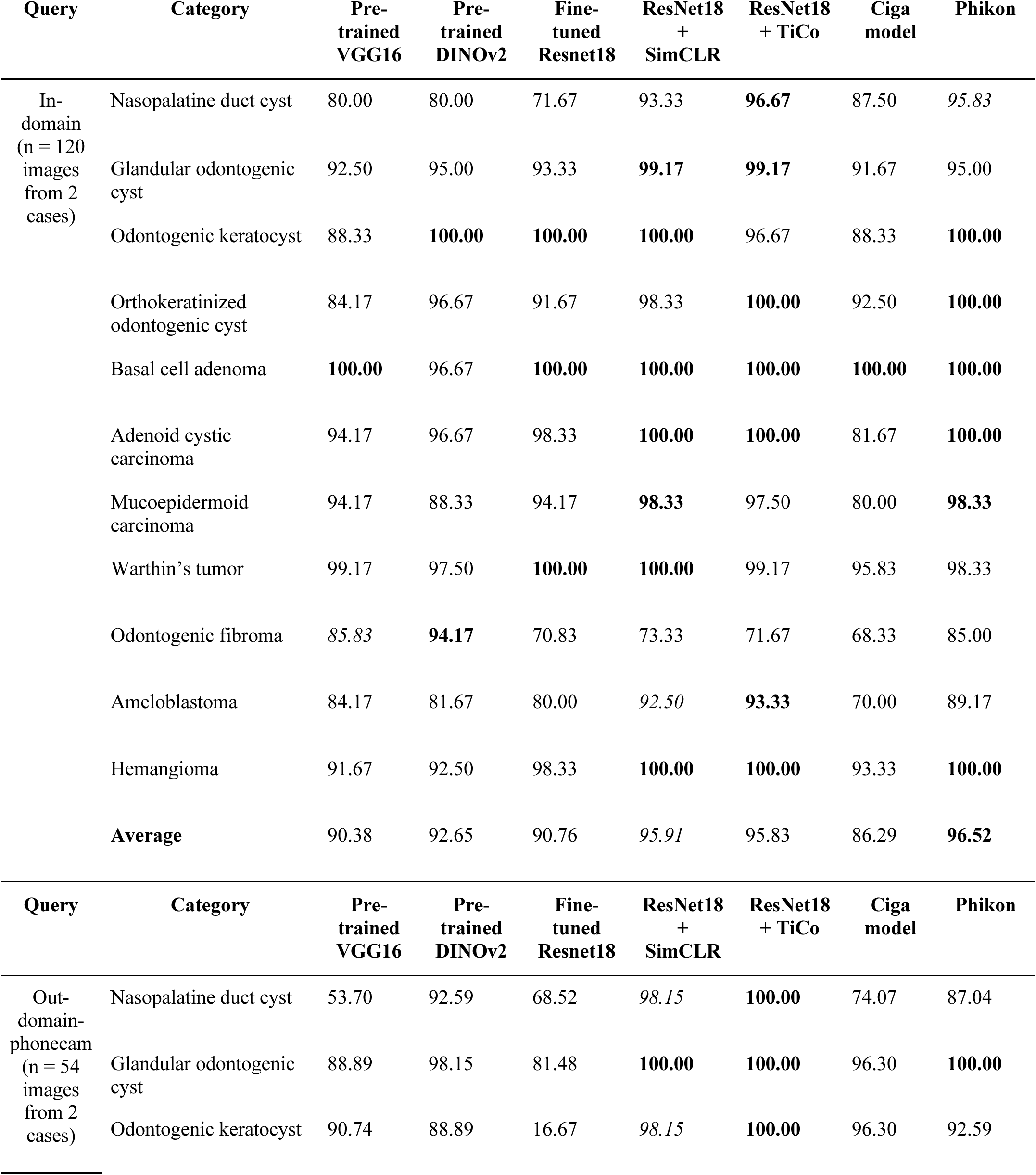
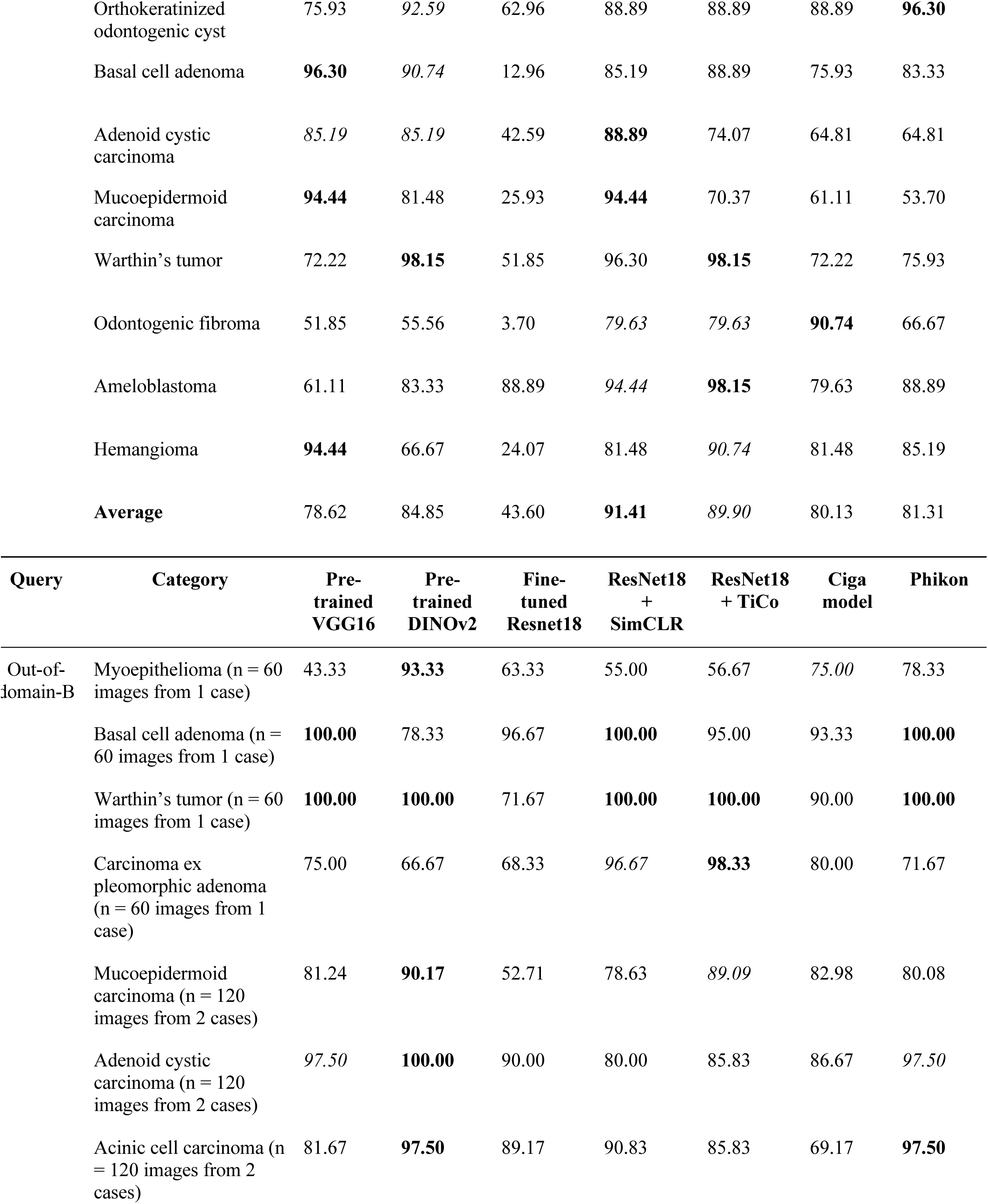
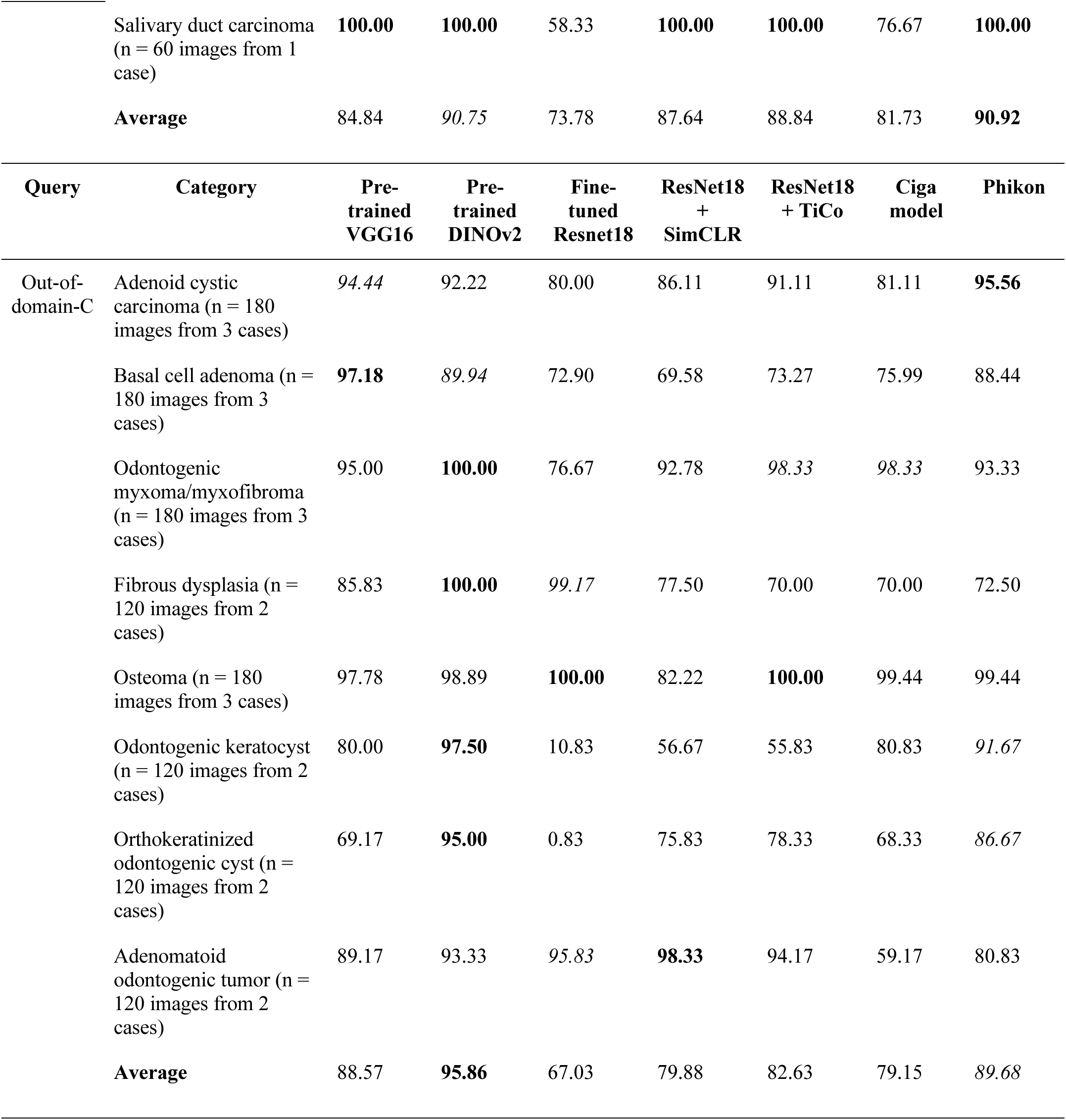
Percentage of queries that retrieved at least one correct diagnosis among the top-10 results for each test query category (%query) in in- and out-of-domain image queries. The performance of the SSL models was superior in most categories. The highest %query for each category is marked in **bold**, and the second highest is in *italics.* (%).

| Query | Category | Pre-trained<br>VGG16 | Pre-trained<br>DINOv2 | Fine-tuned<br>Resnet18 | ResNet18<br>+<br>SimCLR | ResNet18<br>+ TiCo | Ciga<br>model | Phikon |
| --- | --- | --- | --- | --- | --- | --- | --- | --- |
| In-domain<br>(n = 120<br>images<br>from 2<br>cases) | Nasopalatine duct cyst | 80.00 | 80.00 | 71.67 | 93.33 | **96.67** | 87.50 | 95.83 |
|  | Glandular odontogenic cyst | 92.50 | 95.00 | 93.33 | **99.17** | **99.17** | 91.67 | 95.00 |
|  | Odontogenic keratocyst | 88.33 | **100.00** | **100.00** | **100.00** | 96.67 | 88.33 | **100.00** |
|  | Orthokeratinized odontogenic cyst | 84.17 | 96.67 | 91.67 | 98.33 | **100.00** | 92.50 | **100.00** |
|  | Basal cell adenoma | **100.00** | 96.67 | **100.00** | **100.00** | **100.00** | **100.00** | **100.00** |
|  | Adenoid cystic carcinoma | 94.17 | 96.67 | 98.33 | **100.00** | **100.00** | 81.67 | **100.00** |
|  | Mucoepidermoid carcinoma | 94.17 | 88.33 | 94.17 | **98.33** | 97.50 | 80.00 | **98.33** |
|  | Warthin's tumor | 99.17 | 97.50 | **100.00** | **100.00** | 99.17 | 95.83 | 98.33 |
|  | Odontogenic fibroma | 85.83 | **94.17** | 70.83 | 73.33 | 71.67 | 68.33 | 85.00 |
|  | Ameloblastoma | 84.17 | 81.67 | 80.00 | 92.50 | **93.33** | 70.00 | 89.17 |
|  | Hemangioma | 91.67 | 92.50 | 98.33 | **100.00** | **100.00** | 93.33 | **100.00** |
|  | **Average** | 90.38 | 92.65 | 90.76 | 95.91 | 95.83 | 86.29 | **96.52** |
| Query | Category | Pre-trained<br>VGG16 | Pre-trained<br>DINOv2 | Fine-tuned<br>Resnet18 | ResNet18<br>+<br>SimCLR | ResNet18<br>+ TiCo | Ciga<br>model | Phikon |
| Out-domain-phonecam<br>(n = 54<br>images<br>from 2<br>cases) | Nasopalatine duct cyst | 53.70 | 92.59 | 68.52 | 98.15 | **100.00** | 74.07 | 87.04 |
|  | Glandular odontogenic cyst | 88.89 | 98.15 | 81.48 | **100.00** | **100.00** | 96.30 | **100.00** |
|  | Odontogenic keratocyst | 90.74 | 88.89 | 16.67 | 98.15 | **100.00** | 96.30 | 92.59 |

|  |  |  |  |  |  |  |  |
| --- | --- | --- | --- | --- | --- | --- | --- |
| Orthokeratinized odontogenic cyst | 75.93 | 92.59 | 62.96 | 88.89 | 88.89 | 88.89 | **96.30** |
| Basal cell adenoma | **96.30** | 90.74 | 12.96 | 85.19 | 88.89 | 75.93 | 83.33 |
| Adenoid cystic carcinoma | 85.19 | 85.19 | 42.59 | **88.89** | 74.07 | 64.81 | 64.81 |
| Mucoepidermoid carcinoma | **94.44** | 81.48 | 25.93 | **94.44** | 70.37 | 61.11 | 53.70 |
| Warthin’s tumor | 72.22 | **98.15** | 51.85 | 96.30 | **98.15** | 72.22 | 75.93 |
| Odontogenic fibroma | 51.85 | 55.56 | 3.70 | 79.63 | 79.63 | **90.74** | 66.67 |
| Ameloblastoma | 61.11 | 83.33 | 88.89 | 94.44 | **98.15** | 79.63 | 88.89 |
| Hemangioma | **94.44** | 66.67 | 24.07 | 81.48 | 90.74 | 81.48 | 85.19 |
| **Average** | 78.62 | 84.85 | 43.60 | **91.41** | 89.90 | 80.13 | 81.31 |

| Query | Category | Pre-trained VGG16 | Pre-trained DINOv2 | Fine-tuned Resnet18 | ResNet18 + SimCLR | ResNet18 + TiCo | Ciga model | Phikon |
| --- | --- | --- | --- | --- | --- | --- | --- | --- |
| Out-of-domain-B | Myoepithelioma (n = 60 images from 1 case) | 43.33 | **93.33** | 63.33 | 55.00 | 56.67 | 75.00 | 78.33 |
|  | Basal cell adenoma (n = 60 images from 1 case) | **100.00** | 78.33 | 96.67 | **100.00** | 95.00 | 93.33 | **100.00** |
|  | Warthin’s tumor (n = 60 images from 1 case) | **100.00** | **100.00** | 71.67 | **100.00** | **100.00** | 90.00 | **100.00** |
|  | Carcinoma ex pleomorphic adenoma (n = 60 images from 1 case) | 75.00 | 66.67 | 68.33 | 96.67 | **98.33** | 80.00 | 71.67 |
|  | Mucoepidermoid carcinoma (n = 120 images from 2 cases) | 81.24 | **90.17** | 52.71 | 78.63 | 89.09 | 82.98 | 80.08 |
|  | Adenoid cystic carcinoma (n = 120 images from 2 cases) | 97.50 | **100.00** | 90.00 | 80.00 | 85.83 | 86.67 | 97.50 |
|  | Acinic cell carcinoma (n = 120 images from 2 cases) | 81.67 | **97.50** | 89.17 | 90.83 | 85.83 | 69.17 | **97.50** |

|  | Salivary duct carcinoma (n = 60 images from 1 case) | **100.00** | **100.00** | 58.33 | **100.00** | **100.00** | 76.67 | **100.00** |
| --- | --- | --- | --- | --- | --- | --- | --- | --- |
|  | **Average** | 84.84 | *90.75* | 73.78 | 87.64 | 88.84 | 81.73 | **90.92** |
| Query | Category | Pre-trained VGG16 | Pre-trained DINOv2 | Fine-tuned Resnet18 | ResNet18 + SimCLR | ResNet18 + TiCo | Ciga model | Phikon |
| Out-of-domain-C | Adenoid cystic carcinoma (n = 180 images from 3 cases) | *94.44* | 92.22 | 80.00 | 86.11 | 91.11 | 81.11 | **95.56** |
|  | Basal cell adenoma (n = 180 images from 3 cases) | **97.18** | *89.94* | 72.90 | 69.58 | 73.27 | 75.99 | 88.44 |
|  | Odontogenic myxoma/myxofibroma (n = 180 images from 3 cases) | 95.00 | **100.00** | 76.67 | 92.78 | *98.33* | *98.33* | 93.33 |
|  | Fibrous dysplasia (n = 120 images from 2 cases) | 85.83 | **100.00** | *99.17* | 77.50 | 70.00 | 70.00 | 72.50 |
|  | Osteoma (n = 180 images from 3 cases) | 97.78 | 98.89 | **100.00** | 82.22 | **100.00** | 99.44 | 99.44 |
|  | Odontogenic keratocyst (n = 120 images from 2 cases) | 80.00 | **97.50** | 10.83 | 56.67 | 55.83 | 80.83 | *91.67* |
|  | Orthokeratinized odontogenic cyst (n = 120 images from 2 cases) | 69.17 | **95.00** | 0.83 | 75.83 | 78.33 | 68.33 | *86.67* |
|  | Adenomatoid odontogenic tumor (n = 120 images from 2 cases) | 89.17 | 93.33 | *95.83* | **98.33** | 94.17 | 59.17 | 80.83 |
|  | **Average** | 88.57 | **95.86** | 67.03 | 79.88 | 82.63 | 79.15 | *89.68* |

The accuracy calculation excluded the histologic similarity between the query and retrieved images, which provides additional information about the histologic features during diagnosis. To verify whether the SSL models retrieved histologically similar images despite the low MeanAcc, MeanHI was introduced. MeanHI excludes accurate diagnosis and differential diagnosis categories, which are similar to the query and include other inaccurate categories. The lowest overall inaccuracy was consistently achieved by the SSL models, except for the high-magnification in-domain queries, indicating that these models are best at retrieving the most histologically similar images beyond accurate diagnosis (Figures 7C, 7D, Table 6). The top-10 retrieved images of representative cases found by all the tested models are shown in Figures 8 and 9.

**Figure 8:**
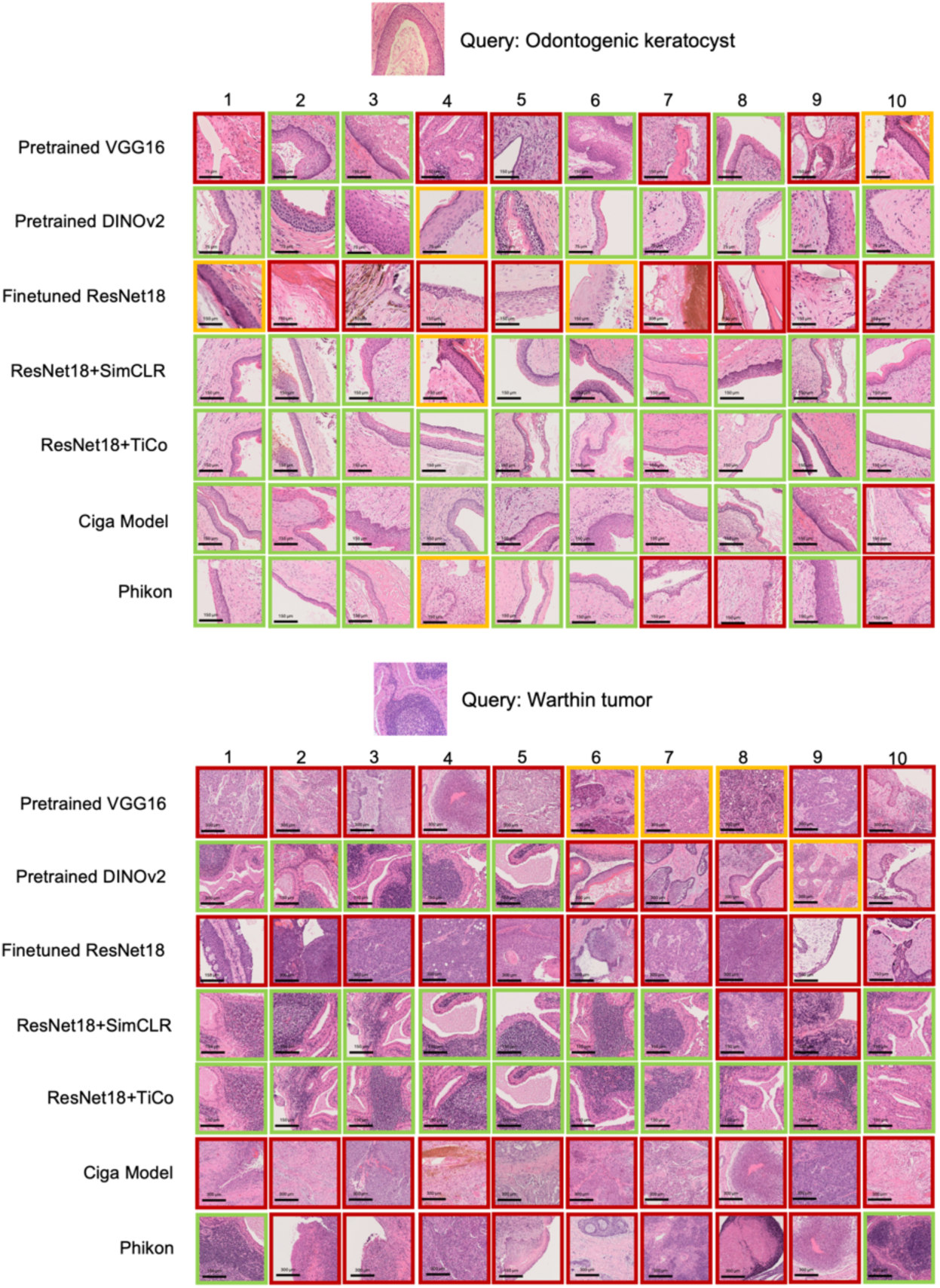
Comparison of the top-10 results of all models for out-of-domain-phonecam queries from different categories. ResNet18+SimCLR and ResNet18+TiCo are consistent with the result that provides the highest Acc for different query categories. More examples are presented in Figure 9 (Green outline: accurate diagnosis category; Yellow outline: differential diagnosis categories; Red outline: inaccurate diagnosis category)

**Figure 9:**
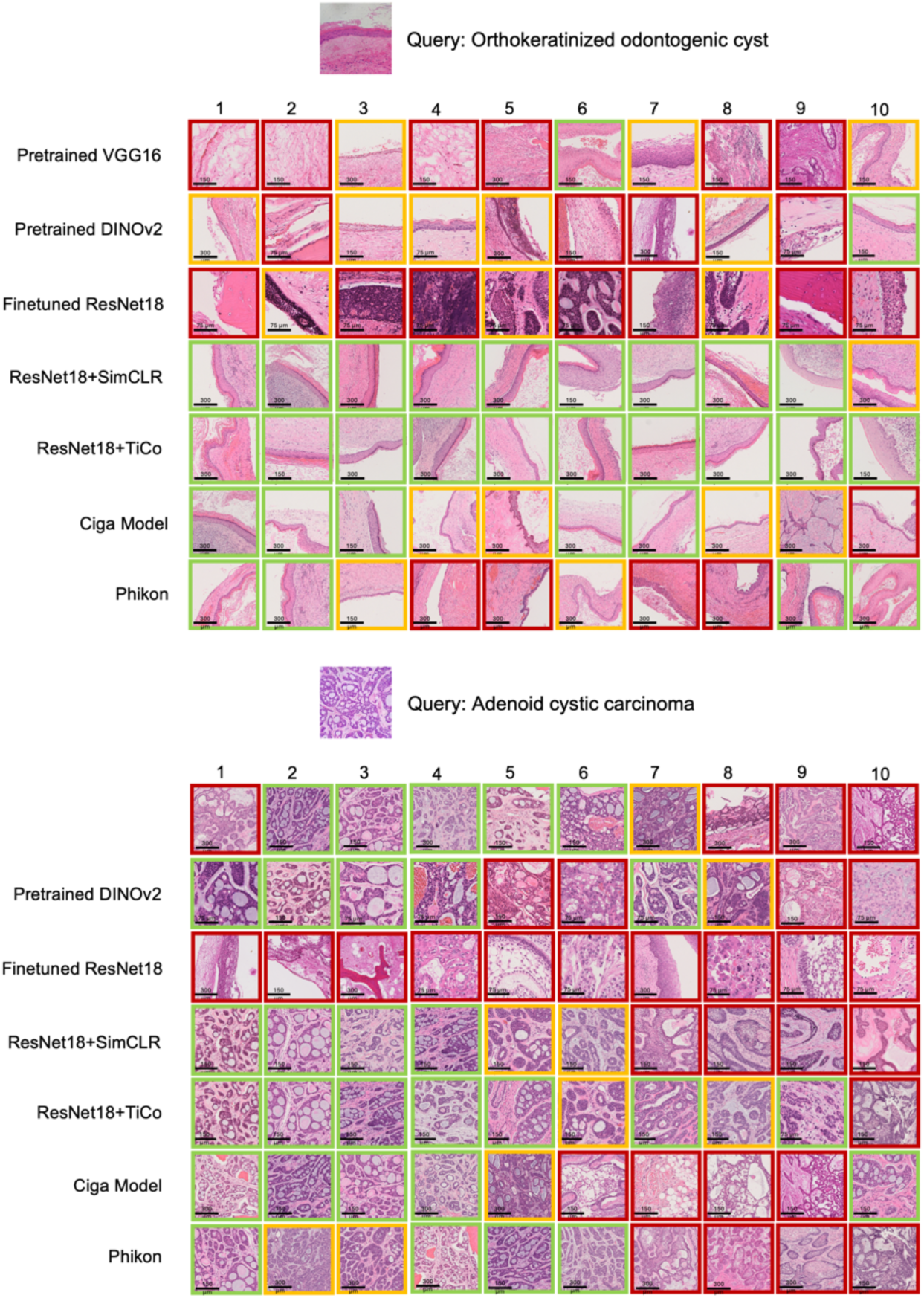
Further examples of comparisons of the top 10 results of all models for out-of-domain-phonecam queries from different categories. ResNet18+SimCLR and ResNet18+TiCo are consistent with the result providing the highest Acc for different query categories. The categories of retrieved images belonging to the differential diagnoses show which retrieved images have histologic similarity to the query. This comparison demonstrates the SSL model’s capability to retrieve histologically similar images when the exact accurate diagnosis is not retrieved. The pre-trained DINOv2 and Phikon also show such potential albeit less consistently across the query category than the SSL models. (Green outline: accurate diagnosis category; Yellow outline: differential diagnosis categories; Red outline: inaccurate diagnosis categories)

**Table 6.**
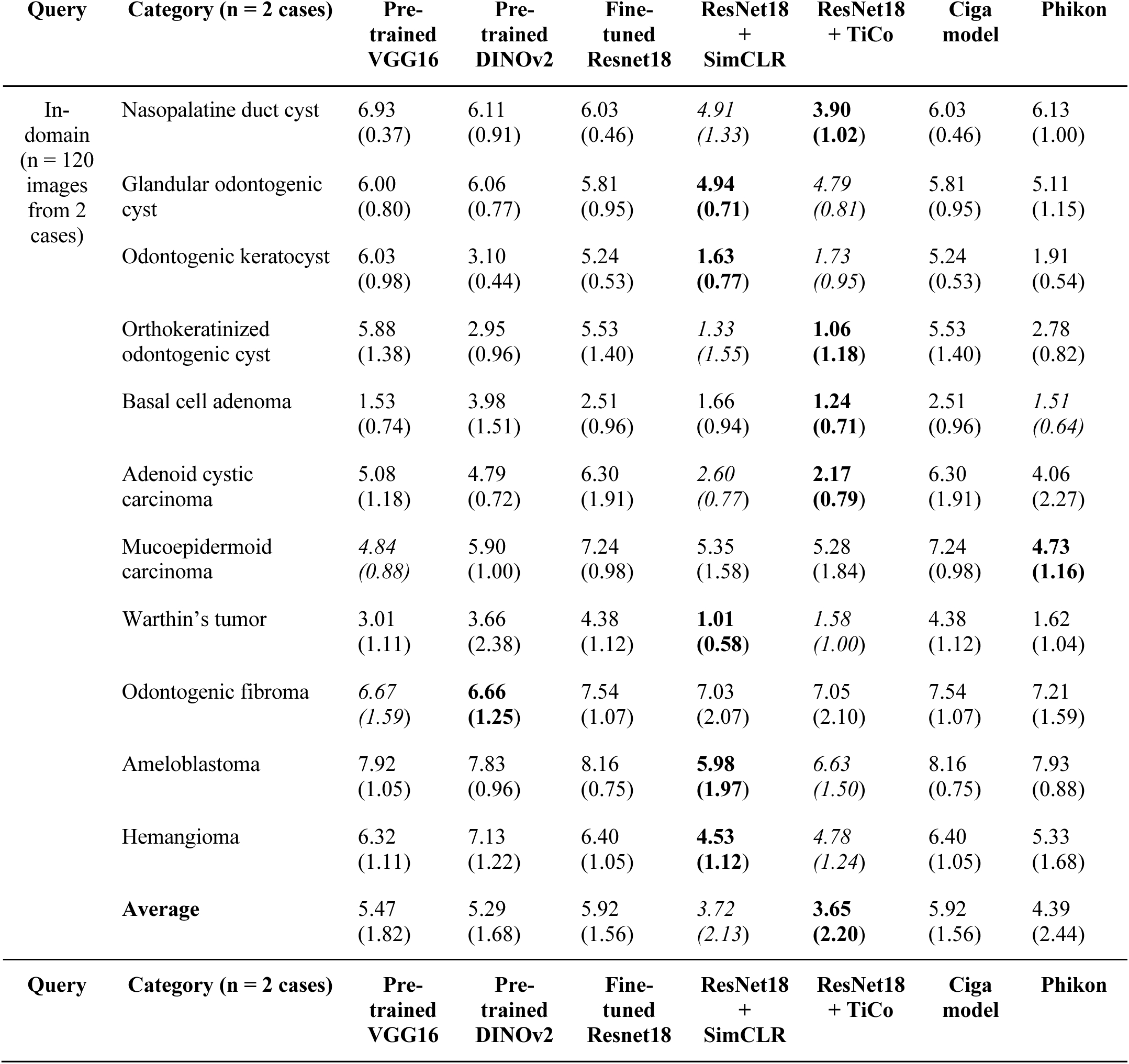
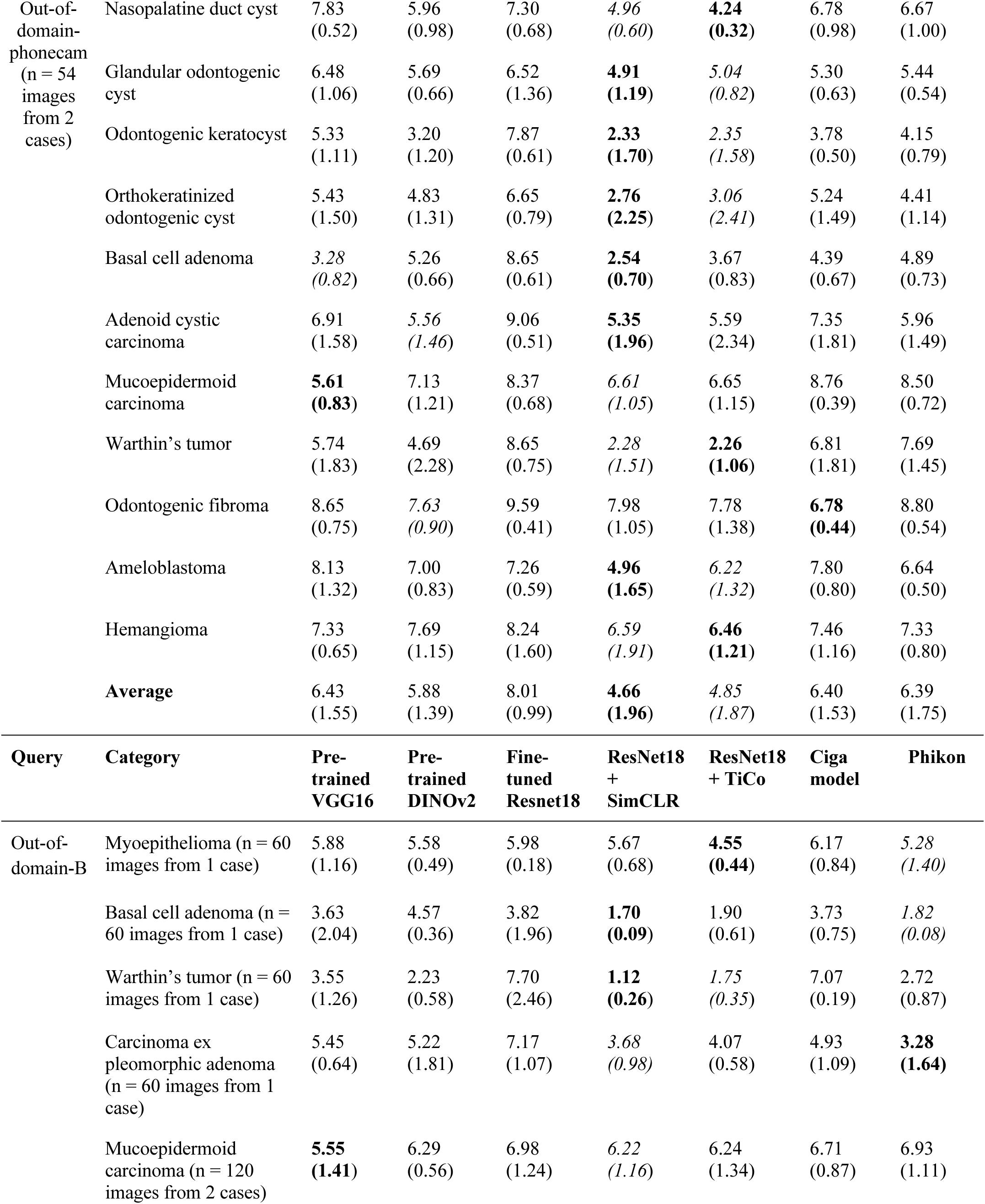
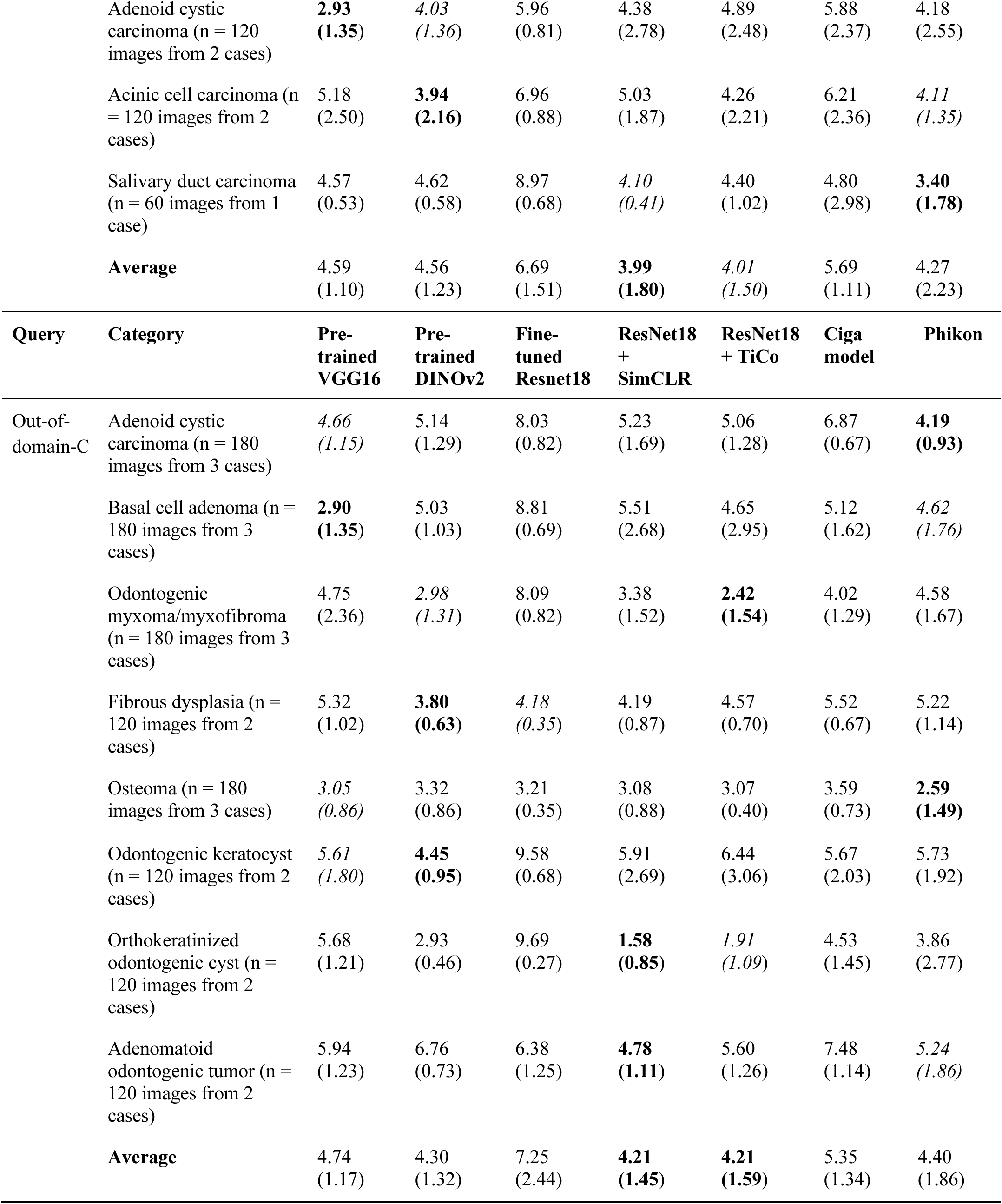
MeanHI (SD) for each test query category for in- and out-of-domain image queries. The best histologic similarity was consistently retrieved by the SSL models. The histologic similarity of the results was consistently best retrieved by the SSL models. It is noteworthy that a lower MeanHI is preferable. The lowest MeanHI for each category is marked in **bold** and the second lowest is in *italics*.

| Query | Category (n = 2 cases) | Pre-trained<br>VGG16 | Pre-trained<br>DINOv2 | Fine-tuned<br>Resnet18 | ResNet18<br>+<br>SimCLR | ResNet18<br>+ TiCo | Ciga<br>model | Phikon |
| --- | --- | --- | --- | --- | --- | --- | --- | --- |
| In-domain<br>(n = 120<br>images<br>from 2<br>cases) | Nasopalatine duct cyst | 6.93<br>(0.37) | 6.11<br>(0.91) | 6.03<br>(0.46) | *4.91*<br>(1.33) | **3.90**<br>(1.02) | 6.03<br>(0.46) | 6.13<br>(1.00) |
|  | Glandular odontogenic cyst | 6.00<br>(0.80) | 6.06<br>(0.77) | 5.81<br>(0.95) | **4.94**<br>(0.71) | *4.79*<br>(0.81) | 5.81<br>(0.95) | 5.11<br>(1.15) |
|  | Odontogenic keratocyst | 6.03<br>(0.98) | 3.10<br>(0.44) | 5.24<br>(0.53) | **1.63**<br>(0.77) | *1.73*<br>(0.95) | 5.24<br>(0.53) | 1.91<br>(0.54) |
|  | Orthokeratinized odontogenic cyst | 5.88<br>(1.38) | 2.95<br>(0.96) | 5.53<br>(1.40) | *1.33*<br>(1.55) | **1.06**<br>(1.18) | 5.53<br>(1.40) | 2.78<br>(0.82) |
|  | Basal cell adenoma | 1.53<br>(0.74) | 3.98<br>(1.51) | 2.51<br>(0.96) | 1.66<br>(0.94) | **1.24**<br>(0.71) | 2.51<br>(0.96) | *1.51*<br>(0.64) |
|  | Adenoid cystic carcinoma | 5.08<br>(1.18) | 4.79<br>(0.72) | 6.30<br>(1.91) | *2.60*<br>(0.77) | **2.17**<br>(0.79) | 6.30<br>(1.91) | 4.06<br>(2.27) |
|  | Mucoepidermoid carcinoma | *4.84*<br>(0.88) | 5.90<br>(1.00) | 7.24<br>(0.98) | 5.35<br>(1.58) | 5.28<br>(1.84) | 7.24<br>(0.98) | **4.73**<br>(1.16) |
|  | Warthin's tumor | 3.01<br>(1.11) | 3.66<br>(2.38) | 4.38<br>(1.12) | **1.01**<br>(0.58) | *1.58*<br>(1.00) | 4.38<br>(1.12) | 1.62<br>(1.04) |
|  | Odontogenic fibroma | *6.67*<br>(1.59) | **6.66**<br>(1.25) | 7.54<br>(1.07) | 7.03<br>(2.07) | 7.05<br>(2.10) | 7.54<br>(1.07) | 7.21<br>(1.59) |
|  | Ameloblastoma | 7.92<br>(1.05) | 7.83<br>(0.96) | 8.16<br>(0.75) | **5.98**<br>(1.97) | *6.63*<br>(1.50) | 8.16<br>(0.75) | 7.93<br>(0.88) |
|  | Hemangioma | 6.32<br>(1.11) | 7.13<br>(1.22) | 6.40<br>(1.05) | **4.53**<br>(1.12) | *4.78*<br>(1.24) | 6.40<br>(1.05) | 5.33<br>(1.68) |
|  | **Average** | 5.47<br>(1.82) | 5.29<br>(1.68) | 5.92<br>(1.56) | 3.72<br>(2.13) | **3.65**<br>(2.20) | 5.92<br>(1.56) | 4.39<br>(2.44) |
| Query | Category (n = 2 cases) | Pre-trained<br>VGG16 | Pre-trained<br>DINOv2 | Fine-tuned<br>Resnet18 | ResNet18<br>+<br>SimCLR | ResNet18<br>+ TiCo | Ciga<br>model | Phikon |

| Out-of-domain-phonecam<br>(n = 54 images from 2 cases) | Nasopalatine duct cyst | 7.83<br>(0.52) | 5.96<br>(0.98) | 7.30<br>(0.68) | 4.96<br>(0.60) | **4.24**<br>**(0.32)** | 6.78<br>(0.98) | 6.67<br>(1.00) |
| --- | --- | --- | --- | --- | --- | --- | --- | --- |
|  | Glandular odontogenic cyst | 6.48<br>(1.06) | 5.69<br>(0.66) | 6.52<br>(1.36) | **4.91**<br>**(1.19)** | 5.04<br>(0.82) | 5.30<br>(0.63) | 5.44<br>(0.54) |
|  | Odontogenic keratocyst | 5.33<br>(1.11) | 3.20<br>(1.20) | 7.87<br>(0.61) | **2.33**<br>**(1.70)** | 2.35<br>(1.58) | 3.78<br>(0.50) | 4.15<br>(0.79) |
|  | Orthokeratinized odontogenic cyst | 5.43<br>(1.50) | 4.83<br>(1.31) | 6.65<br>(0.79) | **2.76**<br>**(2.25)** | 3.06<br>(2.41) | 5.24<br>(1.49) | 4.41<br>(1.14) |
|  | Basal cell adenoma | 3.28<br>(0.82) | 5.26<br>(0.66) | 8.65<br>(0.61) | **2.54**<br>**(0.70)** | 3.67<br>(0.83) | 4.39<br>(0.67) | 4.89<br>(0.73) |
|  | Adenoid cystic carcinoma | 6.91<br>(1.58) | 5.56<br>(1.46) | 9.06<br>(0.51) | **5.35**<br>**(1.96)** | 5.59<br>(2.34) | 7.35<br>(1.81) | 5.96<br>(1.49) |
|  | Mucoepidermoid carcinoma | **5.61**<br>**(0.83)** | 7.13<br>(1.21) | 8.37<br>(0.68) | 6.61<br>(1.05) | 6.65<br>(1.15) | 8.76<br>(0.39) | 8.50<br>(0.72) |
|  | Warthin’s tumor | 5.74<br>(1.83) | 4.69<br>(2.28) | 8.65<br>(0.75) | 2.28<br>(1.51) | **2.26**<br>**(1.06)** | 6.81<br>(1.81) | 7.69<br>(1.45) |
|  | Odontogenic fibroma | 8.65<br>(0.75) | 7.63<br>(0.90) | 9.59<br>(0.41) | 7.98<br>(1.05) | 7.78<br>(1.38) | **6.78**<br>**(0.44)** | 8.80<br>(0.54) |
|  | Ameloblastoma | 8.13<br>(1.32) | 7.00<br>(0.83) | 7.26<br>(0.59) | **4.96**<br>**(1.65)** | 6.22<br>(1.32) | 7.80<br>(0.80) | 6.64<br>(0.50) |
|  | Hemangioma | 7.33<br>(0.65) | 7.69<br>(1.15) | 8.24<br>(1.60) | 6.59<br>(1.91) | **6.46**<br>**(1.21)** | 7.46<br>(1.16) | 7.33<br>(0.80) |
|  | **Average** | 6.43<br>(1.55) | 5.88<br>(1.39) | 8.01<br>(0.99) | **4.66**<br>**(1.96)** | 4.85<br>(1.87) | 6.40<br>(1.53) | 6.39<br>(1.75) |
| Query | Category | Pre-trained VGG16 | Pre-trained DINOv2 | Fine-tuned Resnet18 | ResNet18 + SimCLR | ResNet18 + TiCo | Ciga model | Phikon |
| Out-of-domain-B | Myoepithelioma (n = 60 images from 1 case) | 5.88<br>(1.16) | 5.58<br>(0.49) | 5.98<br>(0.18) | 5.67<br>(0.68) | **4.55**<br>**(0.44)** | 6.17<br>(0.84) | 5.28<br>(1.40) |
|  | Basal cell adenoma (n = 60 images from 1 case) | 3.63<br>(2.04) | 4.57<br>(0.36) | 3.82<br>(1.96) | **1.70**<br>**(0.09)** | 1.90<br>(0.61) | 3.73<br>(0.75) | 1.82<br>(0.08) |
|  | Warthin’s tumor (n = 60 images from 1 case) | 3.55<br>(1.26) | 2.23<br>(0.58) | 7.70<br>(2.46) | **1.12**<br>**(0.26)** | 1.75<br>(0.35) | 7.07<br>(0.19) | 2.72<br>(0.87) |
|  | Carcinoma ex pleomorphic adenoma (n = 60 images from 1 case) | 5.45<br>(0.64) | 5.22<br>(1.81) | 7.17<br>(1.07) | 3.68<br>(0.98) | 4.07<br>(0.58) | 4.93<br>(1.09) | **3.28**<br>**(1.64)** |
|  | Mucoepidermoid carcinoma (n = 120 images from 2 cases) | **5.55**<br>**(1.41)** | 6.29<br>(0.56) | 6.98<br>(1.24) | 6.22<br>(1.16) | 6.24<br>(1.34) | 6.71<br>(0.87) | 6.93<br>(1.11) |

|  |  |  |  |  |  |  |  |
| --- | --- | --- | --- | --- | --- | --- | --- |
| Adenoid cystic carcinoma (n = 120 images from 2 cases) | **2.93**<br>**(1.35)** | *4.03*<br>*(1.36)* | 5.96<br>(0.81) | 4.38<br>(2.78) | 4.89<br>(2.48) | 5.88<br>(2.37) | 4.18<br>(2.55) |
| Acinic cell carcinoma (n = 120 images from 2 cases) | 5.18<br>(2.50) | **3.94**<br>**(2.16)** | 6.96<br>(0.88) | 5.03<br>(1.87) | 4.26<br>(2.21) | 6.21<br>(2.36) | *4.11*<br>*(1.35)* |
| Salivary duct carcinoma (n = 60 images from 1 case) | 4.57<br>(0.53) | 4.62<br>(0.58) | 8.97<br>(0.68) | *4.10*<br>*(0.41)* | 4.40<br>(1.02) | 4.80<br>(2.98) | **3.40**<br>**(1.78)** |
| **Average** | 4.59<br>(1.10) | 4.56<br>(1.23) | 6.69<br>(1.51) | **3.99**<br>**(1.80)** | *4.01*<br>*(1.50)* | 5.69<br>(1.11) | 4.27<br>(2.23) |

| Query | Category | Pre-trained VGG16 | Pre-trained DINOv2 | Fine-tuned Resnet18 | ResNet18 + SimCLR | ResNet18 + TiCo | Ciga model | Phikon |
| --- | --- | --- | --- | --- | --- | --- | --- | --- |
| Out-of-domain-C | Adenoid cystic carcinoma (n = 180 images from 3 cases) | *4.66*<br>*(1.15)* | 5.14<br>(1.29) | 8.03<br>(0.82) | 5.23<br>(1.69) | 5.06<br>(1.28) | 6.87<br>(0.67) | **4.19**<br>**(0.93)** |
|  | Basal cell adenoma (n = 180 images from 3 cases) | **2.90**<br>**(1.35)** | 5.03<br>(1.03) | 8.81<br>(0.69) | 5.51<br>(2.68) | 4.65<br>(2.95) | 5.12<br>(1.62) | *4.62*<br>*(1.76)* |
|  | Odontogenic myxoma/myxofibroma (n = 180 images from 3 cases) | 4.75<br>(2.36) | *2.98*<br>*(1.31)* | 8.09<br>(0.82) | 3.38<br>(1.52) | **2.42**<br>**(1.54)** | 4.02<br>(1.29) | 4.58<br>(1.67) |
|  | Fibrous dysplasia (n = 120 images from 2 cases) | 5.32<br>(1.02) | **3.80**<br>**(0.63)** | *4.18*<br>*(0.35)* | 4.19<br>(0.87) | 4.57<br>(0.70) | 5.52<br>(0.67) | 5.22<br>(1.14) |
|  | Osteoma (n = 180 images from 3 cases) | *3.05*<br>*(0.86)* | 3.32<br>(0.86) | 3.21<br>(0.35) | 3.08<br>(0.88) | 3.07<br>(0.40) | 3.59<br>(0.73) | **2.59**<br>**(1.49)** |
|  | Odontogenic keratocyst (n = 120 images from 2 cases) | *5.61*<br>*(1.80)* | **4.45**<br>**(0.95)** | 9.58<br>(0.68) | 5.91<br>(2.69) | 6.44<br>(3.06) | 5.67<br>(2.03) | 5.73<br>(1.92) |
|  | Orthokeratinized odontogenic cyst (n = 120 images from 2 cases) | 5.68<br>(1.21) | 2.93<br>(0.46) | 9.69<br>(0.27) | **1.58**<br>**(0.85)** | *1.91*<br>*(1.09)* | 4.53<br>(1.45) | 3.86<br>(2.77) |
|  | Adenomatoid odontogenic tumor (n = 120 images from 2 cases) | 5.94<br>(1.23) | 6.76<br>(0.73) | 6.38<br>(1.25) | **4.78**<br>**(1.11)** | 5.60<br>(1.26) | 7.48<br>(1.14) | *5.24*<br>*(1.86)* |
|  | **Average** | 4.74<br>(1.17) | 4.30<br>(1.32) | 7.25<br>(2.44) | **4.21**<br>**(1.45)** | **4.21**<br>**(1.59)** | 5.35<br>(1.34) | 4.40<br>(1.86) |

## Discussion

The diagnosis of oral pathology has long depended on histopathological image observation, which can be a burden for pathologists, especially when dealing with rare cases. In the last decade, various machine learning methods have been proposed to aid in histopathological diagnosis and improve speed and accuracy to avoid delays in diagnosis. However, the exploration for oral histopathology diagnosis has been hampered by the difficulty of obtaining an adequately extensive database that includes rare cases and constructing an effective model. To our knowledge, this is the first study to construct a large database of 30 oral tumor categories, with an additional 21 categories used as the model-training dataset.

Many have argued that CBIR has greater advantages in this field. Pathologists can review CBIR results to make a final decision. However, decision bias may occur when the algorithm is unreliable. To find the best way to represent images for CBIR, we compared different methods of training the feature encoder. We then ranked the similarity of all images in the database to the test queries. The gradual concept of similarity and the multiple-ranked results of CBIR pose a challenge in interpretation. Four evaluation measures were used: MeanAUC from the whole database similarity rank; MeanAcc, %query, and MeanHI from the top-10 most similar images. MeanAUC assumes all rank cut-points are relevant to model performance in extracting histologic features, while the top-10 similar results are relevant during image observation by pathologists in future CBIR implementation. Our findings suggest that model training for feature extraction using an in-house dataset with SSL methods outperforms other popular methods in retrieving images with an accurate diagnosis and similar histology: in-domain queries in 73% of categories and out-of-domain-phonecam queries in 64% of categories (Table 3). This was supported by their MeanAcc which was superior in 63% of in-domain query categories and 82% of out-of-domain-phonecam query categories (Table 4). There was no significant difference in MeanAcc between the SimCLR and TiCo models for both query categories. The MeanAcc superiority of the SSL models was consistent at the in-domain low and moderate magnification levels (Figure 7A) and at out-of-domain-phonecam at all magnification levels (Figure 7B). Additionally, both SSL models retrieved fewer images without histologic similarity at all magnification levels for both query categories (Figures 7C, 7D), meaning when low accuracy is achieved in the result, the users get several options that are histologically similar upon CBIR implementation with the SSL models because they belong to the textbook differential diagnosis categories. From this result, users could proceed with the additional tests more easily than having to do the preliminary reference search manually. Our dataset had overlapping image patches with similar histologic features. In this situation, the SSL method was superior because it compensated for the lack of a labeled dataset for learning representations that cluster the data during training based on semantic classes in conjunction with convolutional neural networks as feature extractors, regardless of the category [20].

The most impressive performance was shown for Warthin’s tumor query, with MeanAUC values greater than 0.960 in every query set using the SSL models (Table 3). Histologically, Warthin’s tumors consist of varying proportions of papillary cystic structures lined by two layers of oncocytic epithelial cells and a lymphoid stroma with germinal centers. It is one of the most common tumors of the salivary gland, especially the parotid gland, and is generally easy to diagnose microscopically owing to its characteristic pattern [19].

The SLL models were successful for most of the test query categories. Out of those categories, the MeanAcc of the SLL model for the ameloblastoma query was lower (Table 3). Although ameloblastoma is one of the most common odontogenic tumors, it has diverse histologic variants: follicular, plexiform, acanthomatous, granular, basaloid, desmoplastic, or a mixture of these [17]. This diversity requires an adequate representation of each subtype in the database for greater accuracy. However, the %query indicated that the models retrieved a similar ameloblastoma type in the top-10 for more than 93% of the queries tested (Table 5). With several differential diagnoses of ameloblastoma included in the database, the best MeanHI obtained for the in-domain ameloblastoma query was 5.98 by SimCLR (Table 6). These categories can be considered histologically similar only if the characteristics of certain subtypes are captured. For example, islands of odontogenic epithelium with ameloblastic features in the follicular type may resemble ameloblastic fibroma [19]. Updating the database with newly encountered subtypes continuously would improve the accuracy of rare tumor subtypes.

Although CBIR works by retrieving similar images that can be considered a digital second opinion, the result may contain images from different categories, with many having similar or indistinguishable histology. Arguably, the range of MeanAcc values obtained with the SSL models, 1.00 to 8.94 (Tables 3 and 4), is considerably wide. However, 55% to 100% of the total queries retrieved at least one of 10 images from the correct category (Table 5), and 6.46 to 1.12 out of the 10 images had no histologic similarity to the query image (Table 6). This implies that displaying the complete top-10 results, including the correct diagnosis and differential diagnosis, as shown in Figures 8 and 9, could be significant for pathologists to narrow the differential diagnoses and conduct further research efficiently. To further improve usability, it is necessary to include clinical and other findings, such as the location of the tumor, patient history, and diagnostic criteria, which are usually essential to making a diagnosis by pathologists, when developing a CBIR system, especially in the oral region where tissue types are diverse.

This study implements patch-based CBIR. Some CBIR systems can analyze WSIs of which implementation is prospective in developed countries. As expensive WSI scanners are not universally installed in oral laboratories, the image-capturing equipment accessible to pathologists differs considerably across regions. Microscope images captured directly using a smartphone camera could be the easiest mode for education, image sharing, and case consultation [27,28]. By using patch-based CBIR where pathologists only need to select the tumor areas and capture them with smartphone cameras to create input, this technology is more accessible globally. Variations in image color and resolution resulting from these differences hinder obtaining reliable results. We tested the robustness of each model to domain shifts by testing the models on out-of-domain queries using WSIs from multiple institutions captured by different scanners and smartphone cameras. SSL models performed best for most query categories, with SimCLR or TiCo achieving the best MeanAUC for over 68% of out-of-domain query categories, from 0.839 to 0.999 (Table 3), confirming the previous finding that SSL is more robust to domain shifts than supervised learning in some datasets, including pathological images [29]. Interestingly, the performances of the vision transformer models (pre-trained DINOv2 and Phikon) always come second best to the SSL models in MeanAUC and are comparable in MeanAcc to that of the SSL models on out-of-domain query sets (Tables 3 and 4). Although further investigation is needed, this result may be considered when choosing methods for a CBIR system. If the system is designed for an in-house database and query or a scenario applicable in large hospitals, SSL models trained on in-house cases are the optimal choice. However, where the system is designed to handle out-of-domain queries, using a pre-trained vision transformer model becomes a viable alternative to eliminate the need to train the SSL model, which could be computationally expensive.

### Study limitations

Limitations of this study include that the experiments involved a single query to retrieve similar images. An algorithm that supports more information in the query, such as multiple query algorithms and filters for location or other diagnostic criteria, would improve retrieval accuracy and provide better support for diagnosis. Our study is limited to test queries from the same geographical area as the SSL model-training dataset. Collecting query cases from a more diverse area would be beneficial in future CBIR development to further challenge the generalizability of the result. The comparative methodology did not emphasize histopathology characteristics that differentiate between benign or malignant tumors, such as capsule invasion and mitotic activity in basal cell adenoma vs. basal cell adenocarcinoma but focused on how such image retrieval tools would be beneficial in reducing to several differential diagnoses and recalling diagnosis criteria before following up with ancillary tests if necessary. Image retrieval is less likely to mislead decision-makers owing to model overfit than a conventional classification method that predicts the possible tumor diagnosis. Nonetheless, a sequel of observations would still be needed when image retrieval is utilized. This study provides insights as the first step to developing a CBIR algorithm by observing retrieval accuracy with strict tumor category criteria on a relatively small database and did not investigate the impact of the result on decision-making in clinical settings. The implementation of CBIR as a well-rounded system to be incorporated into the comprehensive diagnostic process is beyond the scope of this study and observation of the interaction between pathologists and a CBIR system for common and rare diagnoses is needed before the system is used in clinical settings.

## Conclusion

This study highlighted various methods to develop an effective CBIR model and presented key measures to determine the best approach for future clinical usage. We have shown that using SSL methods for deep neural network training is an effective way to develop a CBIR system for histopathological diagnosis of oral tumors compared to other commonly used methods. Vision transformer models, though slightly less effective than SSL models, still provided strong performance and could be a viable alternative for out-of-domain queries. These approaches have considerable potential to create a clinically useful image retrieval system that accelerates the diagnostic process and improves accuracy.

## Data Availability

All data produced in the present study are available upon reasonable request to the authors.

## Author Contributions

R. R. Herdiantoputri: Contributed to the conception, design, data acquisition, drafting, and critical revision of the manuscript. D. K. contributed to the conception, design, drafting, and critical revision of the manuscript. M. Ochi: Contributed to the design and critical revision of the manuscript. Y. Fukawa: Contributed to data acquisition and critically revised the manuscript. K. Kayamori: Contributed to data acquisition, and critically revised the manuscript., M. Tsuchiya: Contributed to data acquisition, and critically revised the manuscript. Y. Kikuchi: Contributed to data acquisition and critically revised the manuscript. T. Ushiku: Contributed to the data acquisition and critically revised the manuscript. T. Ikeda: Contributed to the data acquisition and critically revised the manuscript. Ishikawa: Contributed to the conception, design, drafting, and critical revision of the manuscript. All authors provided their final approval and agreed to be accountable for all aspects of this work.

## Acknowledgments

We thank Editage (www.editage.jp) for the English language review. We thank the pathologists who contributed to the image verification.

